# Combinatorial miRNA1a/15b interference drives adult cardiac regeneration

**DOI:** 10.1101/2024.04.10.24305521

**Authors:** Ting Yuan, Meiqian Wu, Chaonan Zhu, Hao Yu, Minh Duc Pham, Katharina Bottermann, Yijie Mao, Mathias Langner, Mirko Peitzsch, Arka Provo Das, Jonathan Ward, Peter Mirtschink, Andreas Zeiher, Stefanie Dimmeler, Jaya Krishnan

## Abstract

**BACKGROUND:** Despite its promise, cardiac regenerative therapy remains clinically elusive due to the difficulty of spatio-temporal control of proliferative induction, and the need to coordinately reprogram multiple regulatory pathways to overcome the strict post-mitotic state of human adult cardiomyocytes. The present study was designed to identify a novel combinatorial miRNA therapy to address this unmet therapeutic need.

**METHODS:** We performed a combinatorial miRNA interference screen specifically targeting cardiac-predominant miRNAs regulating key aspects of cardiomyocyte mitotic induction to cell-cycle completion, including sarcomerogenesis, metabolic and cell-cycle control pathways. Cardiomyocyte proliferation and cardiac function were assessed in human cardiac biopsies, human cardiac tissue mimetics and in mouse disease models.

**RESULTS:** We identified combinatorial interference of miR-1a and miR-15b (LNA-1a/15b) as drivers of adult cardiomyocyte proliferation. Due to miR-1a/15b function on multiple processes modulating adult cardiomyocyte mitosis, its inhibition augmented adult cardiomyocyte cell-cycle completion and daughter cell formation, and improved contractility in *in vitro* 2D and 3D ischemic models, and in a mouse model of ST-segment elevation myocardial infarction (STEMI). Due to the cardiac-restricted pattern of miR-1a/15b expression, this strategy provides a feasible strategy for specific cardiomyocyte proliferative induction with minimal risk of neoplasm formation and off-target toxicity.

**CONCLUSIONS:** Combinatorial miR-1a/15b inhibition drives mitotic re-entry in adult cardiomyocytes and improves cardiac function in response to myocardial infarction. Our data provides a novel and clinically feasible LNA-based anti-miR-1a/15b strategy to attenuate heart failure and highlights an underutilized therapeutic strategy for simultaneous co-regulation of multiple disease pathways through combinatorial miRNA interference.

## Introduction

Despite improvements in interventional and pharmacological therapies, heart failure remains a major cause of death globally. It affects 64 million people worldwide and leads to cardiovascular morbidity and mortality ^1^. Patients with heart failure suffer from major impairments in quality of life and poor long-term prognosis ^2^. Loss of cardiomyocytes is a major hallmark of heart failure, in both congenital, age- and pathologic stress-related heart failure, and is linked to an approximate 25% decrease in cardiomyocytes due to cell death ^3^. However, unlike other tissues that can compensate cell death through proliferative induction, cardiomyocytes are post-mitotic and have negligible capacity for proliferation, regeneration or repair to restore damaged tissue and overcome remodeling and fibrotic processes leading to heart failure and death ^4^. Thus, there remains a high unmet need for novel therapeutics to enhance cardiomyocyte regeneration and thereby reduce morbidity and mortality from heart failure.

In mammals, cardiomyocytes exit cell cycle and cease proliferation in the early post-natal period ^5^. Whilst evidence suggests some capacity for adult cardiomyocytes to re-enter the cell cycle particularly in response to myocardial insult or injury, proliferation rates are minimal with negligible physiologic or clinical benefits ^5–7^. Gene therapy approaches have in recent years been utilized to overcome this proliferative block with varying degrees of success ^8,9^. Problematically, these attempts have entailed catheter- or intracardial injection for orthotropic delivery of therapeutics into the myocardium, which potentially limits the accessibility of such therapeutics for patients due to the complex and invasive nature of delivery ^8^. These invasive measures are necessary to direct therapeutic delivery to the heart and to restrict leakage of proliferation drivers to non-cardiac tissues in order to prevent ectopic neoplasm formation. Adding to these concerns, studies indicate that proliferative induction of adult cardiomyocytes requires coordinate regulation of multiple genes to overcome the many innate barriers to adult cardiomyocyte cell-cycle re-entry and mitotic completion, including sarcomeric structural rigidity, stability and density, its anti-proliferative metabolic state and the fact that adult cardiomyocytes are prone to undergo endomitosis, rather than mitosis ^10^. Thus, the need to simultaneous modulate multiple pathways to induce physiologically relevant proliferative rates has raised additional technical challenges and clinical concerns.

microRNAs (miRNAs) are small non-coding RNAs that promote mRNA degradation and translational inhibition, and have been shown to play key roles in heart disease development and progression to heart failure. This is evidenced from large animal preclinical, and clinical trial data from recent and ongoing trials modulating disease-linked miRNAs in heart disease ^11–13^. miRNAs function by regulating key genes involved in proliferation, hypertrophy, sarcomerogenesis, inflammation, metabolic reprogramming and contractility ^14–16^. Whilst miRNAs can regulate the expression of multiple genes by binding to target transcripts to remodel the cellular transcriptome ^17^, it is less clear if modulation of a single miRNA alone is sufficient to re-program all pathways necessary to effectively induce cell-cycle re-entry and completion of adult cardiomyocytes. Given that we were not able to identify single miRNAs capable of robustly modulating all relevant pro-proliferative pathways in cardiomyocytes, we sought to determine if a combinatorial RNA interference (co-RNAi) strategy would provide the necessary reprogramming impetus for adult cardiomyocyte cell-cycle re-entry and mitotic completion. Although largely unexplored in heart disease therapeutics, co-RNAi strategies are widely and successfully applied in anti-viral and cancer therapy, wherein combinatorial targeting of multiple key genes serves to better overcome phenotypic and genomic evolution to increase therapeutic robustness and efficacy ^18–20^. Considering precedent for successful clinical safety and efficacy of miRNA targeting therapies, we rationalized that discovery of combinatorial targeting of cardiac-predominant miRNAs would serve to confer a degree of cardiac-specific therapeutic targeting and provide a feasible route for clinical translation - but also as means to coordinately reprogram the multiple pathways necessary for clinically effective adult cardiomyocyte regeneration.

To this end, we performed *in silico* expression and pathway connectivity analysis to identify miRNAs predominantly expressed in the heart with the capacity to modulate pro-proliferative pathways in cardiomyocytes. These efforts led to identification of 17 miRNAs, including miR-1 (miR-1a in rodents), miR-15a/b, miR16, miR-27b, miR-29a, miR-34a/b/c, miR-92a, miR-132, miR-133a, miR155, miR-195a, miR-208a, miR497a and miR-873a as potential regulators ^21^. Utilizing a novel combinatorial RNAi screening methodology wherein the effects of single miRNA interference was compared on the background of interference of all other miRNAs, we identified a common core combination of miRNAs whose inhibition induced cardiomyocyte proliferation in mature human and rat 2D and 3D *in vitro* cardiac ischemic models. We tested the physiologic relevance and efficacy of combinatorial inhibition of miR-1a and miR-15b in driving cardiac regeneration in mice subjected to myocardial infarction (MI). miR-1a/15b interference with locked nucleic acid (LNA) anti-miRs significantly augmented cardiomyocyte proliferation, as determined by EdU incorporation, phospho-Histone H3 staining, and Aurora B localization in newly proliferating adult daughter cardiomyocytes. The capacity of miR-1a/15b to augment cardiac regeneration was further reflected by the decrease in fibrosis, improved cardiac contractility and the reduced mortality in MI mice treated with miR-1a/15b LNA, but not in placebo controls. Critically, despite its pro-proliferative effects on adult cardiomyocytes, LNA-anti-miR-1a/15b (LNA-1a/15b) did not induce neoplasm formation or neoplastic metabolic reprogramming in non-cardiac tissue. Taken together, our data may provide a novel and clinically feasible strategy to induce adult cardiac regeneration.

## Methods

All material, data, code, and associated protocols will be made promptly available to readers.

### Animals

C57BL/6J mice were obtained from Janvier (Le Genest Saint-Isle, France). All animals used in this experiment were housed in a temperature-controlled room with a 12-h light-dark cycle and were allowed free access to food and water in agreement with NIH animal care guidelines, §8 German animal protection law, German animal welfare legislation and with the guidelines of the Society of Laboratory Animals (GV-SOLAS) and the Federation of Laboratory Animal Science Associations (FELASA). 12 week-old C57BL/6J mice were subjected to myocardial infarction (MI) surgery, and LNA delivery was performed 4 h after MI. Mice were killed and hearts isolated at the end of experiment. All protocols were approved by the ethics committee of the regional council (Regierungspräsidium Darmstadt, Hesse, Germany).

### Human heart biopsies

Human heart biopsies were provided by Prof. Dr. Silke Kauferstein (Goethe University Hospital, Frankfurt am Main, Germany). The human heart biopsies were conducted in compliance with the local ethics committee. Samples from healthy hearts as control and from hearts macroscopically visible signs of acute cardiac infarction as MI hearts, and total miRNA was extracted.

### Myocardial infarction (MI)

MI was performed in 12-week-old male C57BL/6J mice, and was induced by permanent ligation of left anterior descending coronary artery (LAD). The mice were monitored up to 28 days after surgery, and the heart dimensions and cardiac function were determined by echocardiography on days 0, 7, 14, 21, and 28. 4 hours after MI, *in vivo* miRCURY LNA Power Inhibitors were injected intraperitoneally (i.p.) into mice at a dose 10 mg/kg. The following *in vivo* miRCURY LNA Power Inhibitors were purchased from QIAGEN: LNA-miR-1a-3p (339203 YCI0200659-FZA), LNA-miR-15b-5p (339203 YCI0200641-FZA), and negative control LNA (339203 YCI0200578-FZA).

### Isolation and maintenance of primary neonatal rat cardiomyocytes

Mated female Sprague Dawley rats were obtained from Janvier (Le Genest Saint-Isle, France). Primary neonatal rat cardiomyocytes (NRCs) were isolated from post-natal day 1 (P1) and P7 rat pups by using the neonatal heart dissociation kit (Miltenyi Biotec) and the gentleMACS Dissociator according to the manufacturer’s instruction. Isolated cells were pre-plated in plating medium (DMEM high glucose, M199 EBS (BioConcept), 10% horse serum, 5% fetal calf serum, 2% L-glutamine and 1% penicillin/streptomycin) for 75 min in 10 cm cell culture dishes (Nunc) at 37°C and 5% CO2 at humidified atmosphere to get rid of fibroblasts as described previously ^22^. NRCs were plated on Type-I bovine Collagen-coated (Advanced Biomatrix) plates or dishes in plating medium. 24 h after isolation of NRCs, the plating medium was changed to maintenance medium (DMEM high glucose, M199 EBS, 1% horse serum, 2% L-glutamine and 1% penicillin/streptomycin).

### Isolation and maintenance of primary rat fibroblasts

Primary rat fibroblasts were isolated from P1 rat pups. After pre-plating the cells in 10 cm cell culture dishes for 75 min, the suspension cells were harvested as cardiomyocytes, the attached cells will be digested and harvested as fibroblasts. The fibroblasts were cultured in DMEM medium with 10% FBS, 2% L-glutamine and 1% penicillin/streptomycin at 37°C and 5% CO2 at humidified atmosphere.

### Preparation and maintenance of human iPSC-CMs

Human induced pluripotent stem cells (hiPSCs) were purchased from Cellular Dynamics International (CMC-100-010-001) and cultured as recommended by the manufacturer. The human iPSC-derived cardiomyocytes (hiPSC-CMs) were reprogrammed using the STEMdiff^TM^ Cardiomyocyte Differentiation Kit (STEMCELL Technologies) according to the manufacture’s protocol. Briefly, human iPS cells were plated at cell density of 3.5×10^5^ cells/well on Matrigel coated 12-well-plates using mTeSR^TM^ medium supplemented with 5uM ROCK inhibitor (Y-27632, STEMCELL Technologies) for 24 h. After 1 day (−1), the medium was replaced with fresh TeSR™ medium. To induce cardiac differentiation, the TeSR™ medium was replaced with Medium A (STEMdiff™ Cardiomyocyte Differentiation Basal Medium containing Supplement A) at day 0, Medium B (STEMdiff™ Cardiomyocyte Differentiation Basal Medium containing Supplement B) at day 2, Medium C (STEMdiff™ Cardiomyocyte Differentiation Basal Medium containing Supplement C) at day 4 and day 6. On day 8, medium was switched to STEMdiff™ Cardiomyocyte Maintenance Medium with full medium changes every 2 days, to promote further differentiation into mature cardiomyocyte cells. All experiments were performed in the hiPSC-CMs at day 40. Hypoxic condition was achieved by using the Hypoxia chamber and the hiPSC-CMs were cultured at 1% O2 for 2 days.

### Human cardiac mimetics formation Technique

Triculture cardiac mimetics were created by hiPSC-CMs. Aggrewell^TM^ 800 microwell culture plates were used to create the human cardiac mimetics in STEMdiff™ Cardiomyocyte Support Medium (STEMCELL Technologies). Approximately 900,000 wells in 2 mL cell suspension were pipetted into each well. After 2 days of culture, medium was switched to STEMdiff™ Cardiomyocyte Maintenance Medium for long term culture.

### EdU labeling *in vitro*

The Click-iT™ EdU Cell Proliferation Kit for Imaging, Alexa Fluor™ 488 dye (C10337, Invitrogen,CA, USA), Alexa Fluor™ 647 dye (C10640, Invitrogen, CA, USA) and Click-iT™ Plus EdU Cell Proliferation Kit for Imaging were used in the study according to the manufacturer’s protocol. The NRCs were labeled by 2 μM EdU for 2 days, the rat fibroblasts were labeled by 2 μM EdU for 4 h, and the hiPSC-CMs were labeled by 2 μM EdU for 6 h.

### Contractility measurement

Every single human cardiac mimetic was transferred into 96-well-plate and treated with either LNA-1a/15b or LNA-NC. Hypoxic condition was achieved by using the Hypoxia chamber and cardiac organoids were cultured at 1% O2 for 3 days, then the contractility will be analyzed by IonOptix system. Units/pixels were determined by calibrating the system with a micrometer.

### *In vitro* administration of locked nucleic acids (LNA)

miRCURY LNA Power Inhibitors were directly added to the cell culture medium at a final concentration of 50 nM. All miRCURY LNA Power Inhibitors and negative control LNA were purchased from QIAGEN: LNA-miR-1a-3p (1999990-801), LNA-miR-15a-5p (339146 YCI0200640-FDA), LNA-miR-15b-5p (339146 YCI0200641-FDA), LNA-miR-16 (339146 YCI0200647-FDA), LNA-miR-27b-5p (339146 YCI0200648-FDA), LNA-miR-29a-3p (339146 YCI0200644-FDA), LNA-miR-34a-5p (339146 (YCI0200649-FDA), LNA-miR-34b-5p (339146 YCI0200645-FDA), LNA-miR-34c-5p (339146 YCI0200650-FDA), LNA-miR-92a-3p (339146 YCI0200646-FDA), LNA-miR-132-3p (1999990-801), LNA-miR-133a-3p (1999990-801), LNA-miR-155-3p (1999990-801), LNA-miR-195a-5p (339146 YCI0200642-FDA), LNA-miR-208a-3p (1999990-801), LNA-miR-497a-5p (339146 YCI0200643-FDA), LNA-miR-873a-5p (1999990-801), and negative control LNA CEL-CONTROL-INH (339147 YCI0199066-FFA).

### RNA isolation, reverse transcription and qRT-PCR

Samples were harvested in QIAzol Lysis Reagent (QIAGEN), total miRNAs and total mRNAs were isolated with miRNA mini Kit (QIAGEN) according to the manufacturer’s protocol. To assess mature miRNA levels, 10 ng total miRNAs were reverse transcript into cDNA using Taqman^®^ Advanced miRNA cDNA synthesis kit (Thermo Fisher Scientific) following the manufacturer’s instructions. The Applied Biosystems StepOnePlus Real-Time PCR system (Applied Biosystems, CA, USA) with TaqMan^®^ fast Advanced PCR Master Mix (Thermo Fisher Scientific) were used for analysis. TaqManTM Advanced miRNA Assays for mmu-miR-1a-3p (ID: mmu482914_mir), has-miR-1-3p (ID: mmu482914_mir), rno-miR-15a-5p (ID: rno483022_mir), mmu-miR-15b-5p (ID: mmu482957_mir), rno-miR-16-5p (ID: rno481312_mir), rno-miR-27b-5p (ID: rno481016_mir), hsa-miR-27b-sp (ID: 478789_mir), hsa-miR-29a-3p (ID: 478587_mir), hsa-miR-34a-5p (ID: rno481304_mir), miR-34b-5p (ID: 002617), hsa-miR_34c-5p (ID: 478052_mir), miR-92a-3p (ID: 000430), rno-miR-132-3p (ID: rno480919_mir), rno-miR-133a-3p (ID: rno481491_mir), mmu-miR-155-3p (ID: mmu481328_mir), mmu-miR-195a-5p (ID: mmu482953_mir), miR-208a-3p (ID: 463567-mat), mmu-miR-497a-5p (ID: mmu482607_mir), miR-873a-5p (ID: rno481425_mir), hsa-miR-26a-5p (ID: 477995_mir), and hsa-26a-5p (ID: 000405). 200ng total RNAs were reverse transcript into cDNA using QuantiTect Reverse Transcription Kit (QIAGEN) according to the manufacturer’s protocol. The Applied Biosystems StepOnePlus Real-Time PCR system (Applied Biosystems, CA, USA) with Fast SYBR^®^ Green Master Mix (Thermo Fisher Scientific) were used for analysis. Gene expression levels were normalized against the housekeeping gene *Hprt1.* The following qRT-PCR primers were used: Mouse: *Nppa*: 5’ GGAGGTCAACCCACCTCTGA 3’ and 5’ CGAAGCAGCTGGATCTTCGT 3’; *Myh6*: 5’ CGGTGACAGTGGTAAAGGCA 3’ and 5’ GGAATGATGCAGCGCACAAA 3’; *Myh7*: 5’ AGAAGAAGGTGCGCATGGAC 3’ and 5’ ATCCTGGCATTGAGTGCATTT 3’; *Tgf-β2*: 5’TTGTTCCACAGGGGTTAAGG 3’ and 5’ AGCTCGGTCCTTCA GATCCT 3’; *Col3a1*: 5’ CTGTAACATGGAAACTGGGGAAA 3’ and 5’ CCATAGCTGAACTGAAAACCACC 3’; *Thbs1*: 5’ CACCTCTCCGGGTTACTG AG 3’ and 5’ GCAACAGGAACAGGACACCTA 3’; *Hprt*: 5’ CCACTTGTGACGA AAGCACC 3’ and 5’ GTTGTCTACGCTCTGGCAGT 3’.

### Immunofluorescence staining

Immunofluorescence staining was performed as described previously ^23^. After fixation of cells with 4% paraformaldehyde (PFA)/PBS, the cells were permeabilized and incubated overnight at 4°C with primary antibodies against sarcomeric α-actinin (Sigma Aldrich), Aurora B (Abcam), α-SMA (Abcam), Ki67 (Abcam), pHH3 (Abcam), cleaved (Cl.) Caspase 3 (Abcam) or Tomato lectin (TL) (Sigma Aldrich) diluted in 2% HS/PBS (v/v). After 3 washes with PBS for 5 min, cells were incubated with 4′,6-diamidino-2-phenylindole (DAPI; Thermo Fisher Scientific), AlexaFluor 555 anti-mouse (Thermo Fisher Scientific) and AlexaFluor 488 anti-rabbit (Thermo Fisher Scientific) secondary antibody for 1 h at room temperature. Dishes were mounted onto glass slides (Fisher Scientific) with a drop of ProLong Antifade (Thermo Fisher Scientific). Fluorescent images were acquired with the SP5 confocal microscopy (Leica) using a 40x magnification.

Following the α-SMA, α-actinin, WGA, the cells or sections were stained with EdU. EdU staining was conducted using Click-iT™ EdU imaging kit (Invitrogen, CA), according to the manufacturer’s protocol. Briefly, after incubation with secondary antibody for 1 h, cells were washed with 3XPBS for 5min. The cells were then incubated with a Click-iT™ reaction cocktail for 30 min for EdU staining, then the cells were washed with 3XPBS for 5min. Finally, fluorescent images were acquired with the SP5 confocal microscopy (Leica) using a 40x magnification.

### ASAT and ALAT measurement

The blood was collected in tubes prefilled with EDTA for determination of Aspartate aminotransferase (ASAT) and Alanine aminotransferase (ALAT) in sera from animals in different groups. Sera samples were measured in a Cobas 8000 Analyzer (Roche Diagnostics, Germany) using Roche reagents following the manufacturer’s instructions.

### Picro Sirius Red Staining

Picro sirius red staining was used to determine collagen deposition and fibrosis in heart cryosections. 0.1% Picrosirius Red solution was prepared by solving 0.5 g Sirius Red (Waldeck GmbH) in 500 mL picric acid (Sigma-Aldrich). The cryosections were washed with water and PBS, and then fixed with 4% paraformaldehyde (PFA) for 30min. Then the cryosections were incubated in the 0.1% Picrosirius Red solution for 1 h. After washing two times with acidified water, the sections were dehydrated with 100% ethanol, cleared with xylene and mounted with Pertex (Medite) as previously described^22^.

### Metabolomics sample preparation

5×10^5^ P1 rat cardiomyocytes were cultured per 3 cm dish. 24h after isolation miRCURY LNA Power Inhibitors were added to maintenance medium at a final concentration of 50 nM. The cell culture plates were snap frozen in liquid nitrogen 48 h after miRCURY LNA Power Inhibitors incubation, right after the cells were washed with 75 mM ammonium carbonate (Sigma), adjusted to pH 7.4 with acetic acid. In addition, 20 mg of organ tissues (liver, kidneys, lungs, spleen and kidneys) were homogenized at full speed. Metabolite extraction from NRC and homogenized tissues was done twice with cold extraction buffer (–20°C) containing acetonitrile: methanol: water in a 40:40:20 ratio. After centrifugation at full speed, sample extract supernatants were dried by vacuum-assisted centrifugation.

### Non-targeted metabolomics

Untargeted metabolomics was conducted through metabolite profiling utilizing liquid chromatography high-resolution tandem mass spectrometry (LC-HR-MS/MS). The experimental setup comprised an ultra-performance liquid chromatography system (Aquity I-class; Waters GmbH, Eschborn, Germany) coupled with a quadrupole – time-of-flight mass spectrometer (QToF) equipped with an additional ion mobility spectrometer (IMS) (VION IMS QToF; Waters GmbH, Eschborn, Germany). Sample extract residues were reconstituted in 200 µL aqueous acetonitrile (10% water) containing 0.1% formic acid and transferred to auto sampler glass vials for analysis. A pooled sample, consisting of 20 µL from each individual sample, was prepared separately and analyzed randomly throughout the experiment. Chromatographic separation utilized hydrophilic interaction chromatography (HILIC) with a BEH Amide column (2.1 x 100 mm, 1.7 µm; Waters GmbH, Eschborn, Germany) at 45°C, employing a gradient of mobile phase A (water with 0.1% formic acid) and mobile phase B (acetonitrile with 0.1% formic acid). The mass range covered mass-to-charge ratios between 50 and 1000 Da, with a total scan time set to 0.3 s. Positive and negative electrospray ionization (ESI) in combination with high-definition data acquisition scan mode (HDMSE) were employed, including ion mobility screening, determination of metabolite-specific collision cross-section (CCS) values, accurate mass, and fragment ion mass screenings.

Ion source parameters in positive ESI mode were set to 1kV and 40V for capillary voltage and sample cone voltage, and 120°C and 550°C for source and desolvation temperature. In negative ESI mode, parameters were adjusted to 1.5kV and 40V, and 120°C and 450°C, respectively. Nitrogen was used as cone gas and desolvation gas with flow rates of 50 L/h and 1000 L/h in positive and negative ESI. Leucine-Enkephalin solution injections were performed for mass correction. Data acquisition and raw data processing were carried out using the Waters Unifi software package 2.1.2. Further data processing involved importing Unifi export files into Progenesis QI software (Nonlinear Dynamics, Newcastle upon Tyne, UK) for peak picking, chromatographic alignment, signal deconvolution, data normalization, and experimental design setup.

For metabolite identification, features obtained from untargeted metabolomics screenings were compared with the Human Metabolome Database (HMDB, www.hmdb.ca) using the Progenesis Metascope plugin, applying a precursor mass accuracy of ≤ 10 ppm and a theoretical fragment mass accuracy of ≤ 10 ppm. Additionally, features were compared to in-house data obtained by injecting metabolite standards in neat solution, allowing deviations of retention time and CCS of 0.3 min and 3%, respectively. Metabolic features without a metabolite annotation were excluded, and the remaining features, along with their signal intensities and identifications, were exported to Microsoft Excel for further data processing.

### Bioinformatic analysis

The P1 rat cardiomyocytes were treated with either control LNA-NC or LNA-1a/15b in hypoxia with 1% O2, and then RNAs were extracted for next generation RNA sequencing (RNA-seq) (Novogene). The integrity and purity of the RNA were assessed, and qualified RNA samples were utilized for PCR amplification in order to construct a cDNA library. The cDNA library was sequenced using an Illumina HiSeq™ 150 platform. For RNA sequencing analysis, raw RNA-Seq data (fastq files) were first cleaned using Trimmomatic v0.39 (Trimmomatic: a flexible trimmer for Illumina sequence data). The cleaned reads were then accurately mapped to the Ensembl rat genome (mRatBN7.2) using HISAT 2.2.1, and sequence comparison/mapping format (sam) files were generated ^24^. The Sam files were then sorted by Samtools v1.19.2 to generate binary comparison/mapping format (bam) files, which HTSeq-count v2.0.3 uses to summarize gene level counts (HTSeq-a Python framework for working with high-throughput sequencing data). DESeq2 ^25^ was used to detect differentially expressed genes (DEGs), and the DEGs with the cut-off criterion (*P* value < 0.05 and Log|FC|>1) were considered for further analysis. The BioSample Database (BioProject ID: PRJNA000000) was selected for the data analysis.

### Statistical Analysis

Data are represented as mean and error bars indicate the standard error of the mean (SEM). Two-tailed unpaired Student’s *t*-tests (Excel) or Mann Whitney test were used as indicated in the respective figure legends. P values were determined with Prism 9.0 (GraphPad) and P < 0.05 was considered statistically significant.

## Results

### Combinatorial miRNA interference (co-miRNAi) screen identifies co-operative cardiomyocyte pro-proliferative drivers

Utilizing the microRNA Disease Database to identify miRNAs linked to myocardial infarction (MI) and associated with cardiac anti-proliferative pathways ^21^, we identified 17 miRNAs as potential regulators of adult cardiomyocyte proliferation. These miRNAs are elevated in MI and linked to proliferative pathway suppression ^16,21,26^. All 17 miRNA were expressed at detectable levels in healthy and infarcted human heart biopsies, albeit elevated in the MI patient biopsies (**Fig S1A**). To explore the individual, and potential cooperative or synergistic consequence of combined miRNA suppression, we developed an LNA combinatorial screening assay connecting specific LNAs anti-miRs with their capacity to induce cardiomyocyte proliferation. Starting with an initial set of LNA anti-miRs targeting all 17 miRNAs, we systematically removed one LNA (targeting a specific miRNA) from the pool of 17 LNAs, to determine how exclusion of a single LNA (targeting one miRNA) would affect cardiomyocyte proliferation. By comparing proliferation rates in samples lacking the single LNA anti-miR against the reference control containing all 17 LNA anti-miRs, this assay identifies individual LNAs that function to drive cardiomyocyte proliferation. This strategy and downstream steps of the workflow are depicted in **Figure 1**.

**Figure 1.**
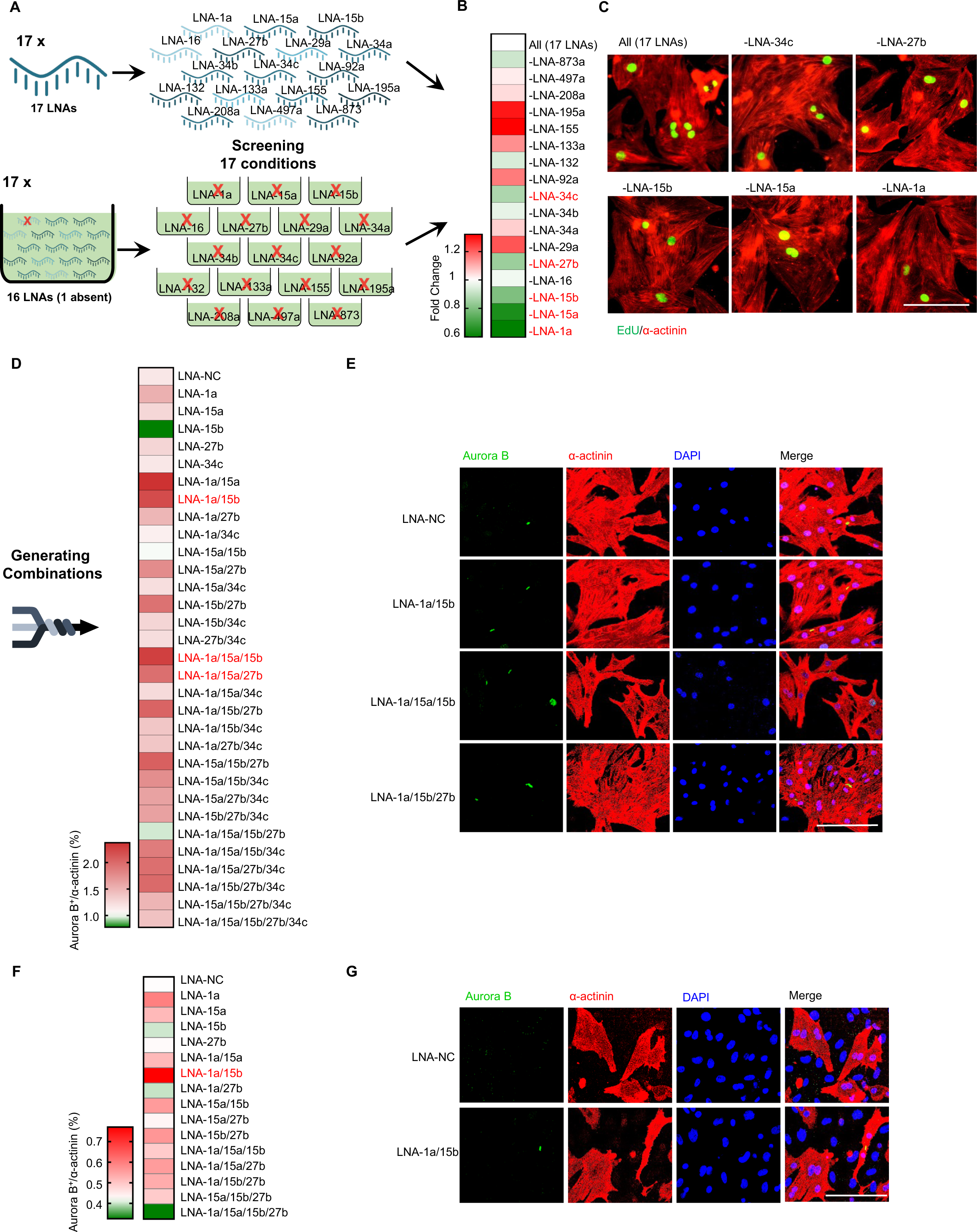
Screening of co-miRNAi that can promote cardiomyocyte proliferation in the P1 and P7 rat cardiomyocytes. (A) Schematic diagram of the research. A set of 17 LNAs have been screened and then all combinations of the top hits will be tested in the neonatal P1 and P7 rat cardiomyocytes. The effects of the best combination will then be validated *in vitro* and *in vivo*. (B, C) Effect of the removal of individual LNA from the pool of 17 LNAs in the neonatal P1 rat cardiomyocytes. Neonatal P1 rat cardiomyocytes were treated with different combinations of LNAs and stained for EdU (green) and cardiomyocyte-specific α-actinin (red). (B) Heat map shows the fold change of EdU^+^ cells in α-actinin^+^ cardiomyocytes. Data are normalized to All (17 LNAs). The red and green colors indicate high and low values, respectively. n=3. (C) Representative images of EdU and α-actinin in cardiomyocytes. (D, E) Neonatal P1 rat cardiomyocytes were treated with different combinations of LNAs for 2 days, and then cardiomyocytes were stained for Aurora B (green), cardiomyocyte-specific α-actinin (red) and DAPI (blue). (D) Heat map shows the quantification of the percentage of Aurora B^+^ cells in α-actinin^+^ cardiomyocytes. The red and green colors indicate high and low values, respectively. (E) Representative images of Aurora B, α-actinin and DAPI in cardiomyocytes. Means of n = 3 biological replicates per group. Scale bar is 100 µm. (F, G) P7 rat cardiomyocytes were treated with different combinations of LNAs for 48 h and stained for Aurora B (green) and α-actinin (red), and DAPI (blue). (F) Heat map shows the quantification of rat α-actinin^+^ cardiomyocytes that were Aurora B^+^ as a percentage of total α-actinin^+^ cells. Data are normalized to LNA-NC. The red and green colors indicate high and low expression values, respectively. n=3. (G) Representative images of Aurora B, α-actinin and DAPI.

We first confirmed that simultaneous interference of all 17 miRNAs by LNA anti-miRs can lead to effective inactivation of all 17 target miRNAs in neonatal P1 rat cardiomyocytes (**Figure S1B)**. Next, we initiated the combinatorial LNA anti-miR screen utilizing 5-Ethynyl-2’-deoxyuridine (EdU)-incorporation as an initial readout for proliferation, with co-staining for the cardiomyocyte specific marker, sarcomeric α-actinin to facilitate identification of cycling cardiomyocytes (**Figure 1B, C**). Strikingly, exclusion of LNA anti-miRs targeting miR-1a, miR-15a, miR-15b, miR-27b and miR-34c led to decreased cardiomyocyte proliferation - indicative of potential cooperative function with other LNA miRNAs in driving cardiomyocyte proliferation.

To confirm these findings, we reverted to screening LNAs targeting miR-1a, miR-15a, miR-15b, miR-27b and miR-34c both individually, and in all possible permutations in P1 rat cardiomyocytes. Whilst EdU serves as an important maker for S-phase DNA replication, it precludes the determination of cell-cycle completion and daughter cell formation ^27^. Hence, we specifically quantified Aurora B staining at telophase as readout for completed cardiomyocyte proliferation (**Figure S2A)**. Telophase is the final phase of mitosis, during which duplicated genetic material carried in the nucleus of a parent cell is separated between two daughter cells. We assessed Aurora B localization at telophase in 31 individual and combinatorial conditions corresponding to the 5 candidate LNA anti-miRs targeting miR-1a, miR-15a, miR-15b, miR-27b and miR-34c. Cardiomyocyte proliferation was assessed by co-localization of contractile ring and midbody Aurora B staining, with sarcomeric α-actinin and DAPI counterstains to mark cardiomyocytes and their nuclei, respectively (**Figure 1D, E**). These analyses revealed that treatment with 3 specific LNA combinations, comprising LNA anti-miRs targeting miR-1a and miR-15b, miR-1a, miR-15a and miR-15b, and miR-1a, miR15a and miR-27b, resulted in consistently increased numbers of cardiomyocytes in telophase compared to control scrambled LNA (LNA-NC) treated cells (**Figure 1D, E; Figure S2B-D**). In accord, all miRNAs targeted within the respective combinations were effectively repressed (**Figure S2E-G**).

The early neonatal mouse heart has some proliferative and regenerative response, but this capacity is lost from P7 ^28^. Thus, to determine the impact of the identified LNA miRNA combinations in post-mitotic cardiomyocytes, all combinations of LNAs targeting miR-1a, miR-15a, miR-15b and miR-27b were screened in P7 rat cardiomyocytes. Utilizing Aurora B staining at telophase as readout, the specific combination of LNAs targeting miR-1a and miR-15b (miR-1a/15b) proved most effective at inducing proliferation in post-mitotic cardiomyocytes (**Figure 1F, G**). Consistent with previous findings, LNAs targeting miR-1a and miR-15b led to efficient target knockdown (**Figure S3**). Thus, combinatorial interference of miR-1a/15b drives post-mitotic cardiomyocyte proliferation and cell-cycle completion in rat cardiomyocytes *in vitro*. To further confirm that the proliferative responses detected as a result of combinatorial inhibition of miR-1a/15b were indeed originating from cardiomyocytes, and from not a hyper proliferative cell-type such as cardiac fibroblasts, we assessed the effects of miR-1a/15b LNA in cardiac fibroblasts. As shown in the **Figure S4A, B**, LNA-anti-miR1a/15b had negligible impact on fibroblasts proliferation. Taken together, these data suggest a degree of cell-type specificity in LNA-miR1a/15b function in proliferative induction.

### Combinatorial miR-1a/15b interference drives adult cardiomyocyte proliferation and maintenance of cardiac function in pathology

The pro-proliferative impact of miR-1a/15b function *in vitro* led us to explore its potential therapeutic utility *in vivo*. To that end, we assessed the effects of LNA-1a/15b treatment in a mouse model of ST-Elevation Myocardial Infarction (STEMI). STEMI is characterized by development of acute myocardial infarction as a result of decreased coronary blood flow, accounting for approximately 40% of all myocardial infarction presentation in hospitals ^29,30^.

We first assessed the efficiency of LNAs targeting miR-1a and miR-15 both individually and in combination in adult mice, as depicted in **Figure S5A**. Both individual and combinatorial delivery of LNAs targeting miR-1a and miR-15b led to effective target suppression (**Figure S5B-D**). Whilst depletion of miR-1a or miR-15b in mouse hearts led to a slight but statistically insignificant increase in proliferation, combinatorial depletion of both miR-1a and miR-15b significantly induced cardiomyocyte proliferation as shown by increased pHH3 (**Figure S5E, F**).

We next examined if inhibition of miR-1a/15b could similarly increase cardiomyocyte proliferation in response to a mouse STEMI model (**Figure 2A**). MI was induced by permanent ligation of left anterior descending coronary artery, resulting in elevated expression of miR-1a and miR-15b (**Figure 2B**). Mice were treated with LNA-1a/15b four hours post-surgery to mimic the average duration that post-STEMI patients receive treatment ^29,30^. At 28 days post-MI surgery the experiment was terminated due to the decline in cardiac function and increased mortality in mice subjected to MI and treated with control non-silencing LNAs (LNA-NC) (**Figure 2C**). LNA-1a/15b treatment effectively suppressed RNA levels of miR-1a and miR-15b in border zone biopsies of the respective mice (**Figure 2D, E**). Echocardiographic measure of ejection fraction (EF), as readout for cardiac contractile function, was rapidly reduced in LNA-NC treated mice subjected to MI concomitant to increased mortality (**Figure 2F, G**). In contrast, mice treated with LNA-1a/15b largely maintained cardiac function and organismal viability despite MI intervention (**Figure 2F, G**). To ascertain the effects of LNA-1a/15b treatment in mice subjected to MI, the heart was first divided into remote and border zones (**Figure 2H**). As the detected rescue and maintenance of healthy cardiac in LNA-1a/15b treated mice would predict for reduced expression of pathologic hypertrophy markers, we examined expression of *atrial natriuretic peptide* (*Nppa*), *beta-myosin heavy chain 6* (*Myh6*) and *beta-myosin heavy chain 7* (*Myh7*) (**Figure 2I**). *Nppa* and *Myh7* RNA levels were reduced after LNA-1a/15b injection in the border zone compared to the control LNA-NC injection (**Figure 2I**). Similarly, MI-induced *Myh7*:*Myh6* ratio was reduced by combinatorial inhibition of miR-1a and miR-15b, indicative of suppression of the pathologic hypertrophic gene program induction in LNA-1a/15b treated hearts (**Figure 2I**). The remote zone serves as an internal tissue control due to its distance from the infarct site, and is thus less affected by tissue damage caused by MI (**Figure 2H**). Consistent with this, LNA-1a/15b treatment primarily impacted hypertrophic marker expression in border zone biopsies, but not in the largely unaffected remote zone (**Figure 2I**).

**Figure 2.**
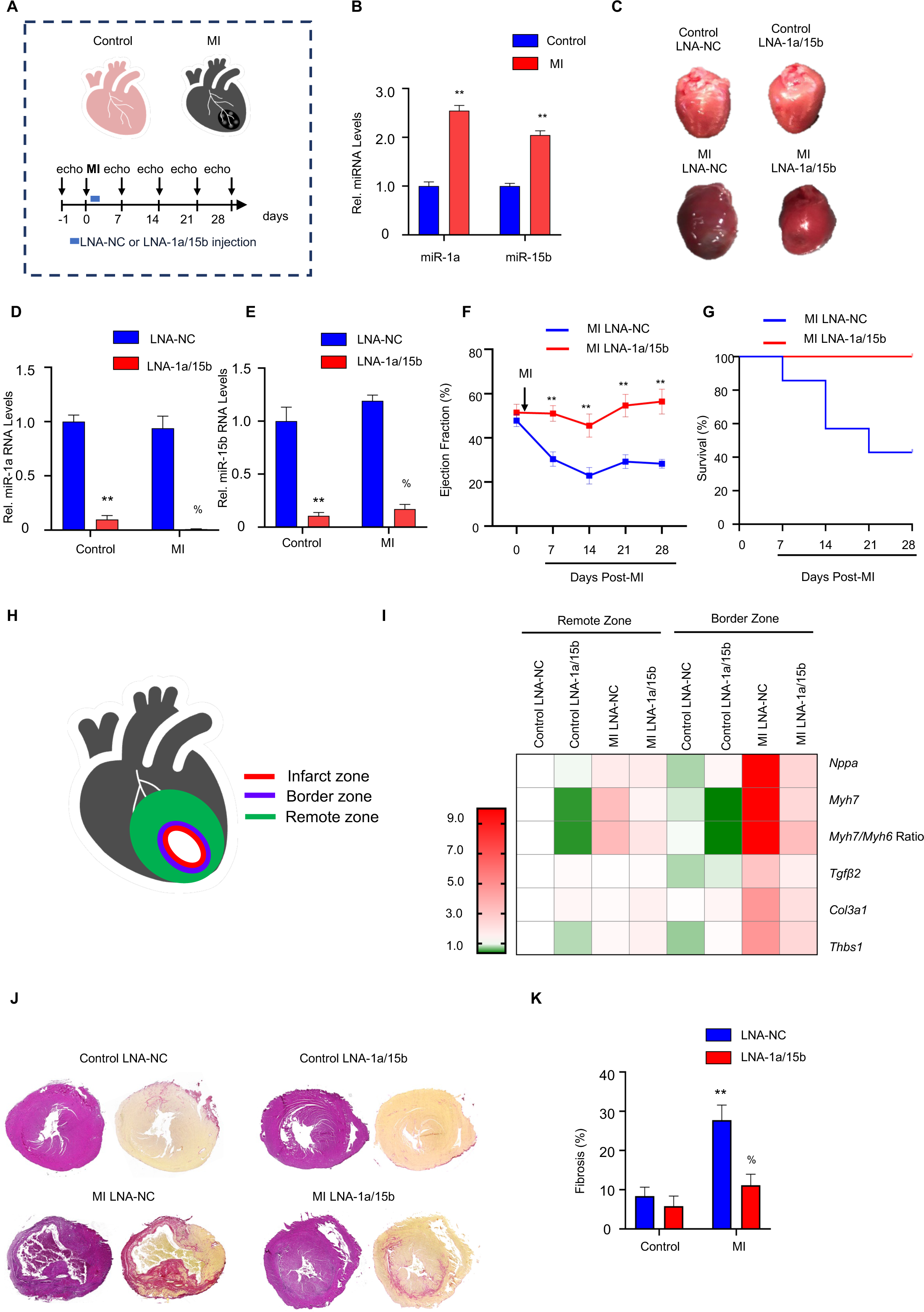
The effects of combinatorial miR-1a/15b interference on myocardial infarction-induced cardiac damage *in vivo*. (A) Schematic representation of the experimental timeline of C57BL/6J mice subjected to MI surgery. The LNA-1a/15b or LNA-NC was injected at 4 h after MI surgery, and echocardiography measurement was performed at day 0, 7, 14, 21 and 28. The mouse hearts were harvested at day 28 after MI surgery. (B) Relative miRNA expression in heart border zone biopsies from mice with MI and healthy controls. Data are expressed as means ± SEM; n=3 mice per group; **P < 0.01 vs. control. Two-tailed unpaired t-test. (C) Whole hearts harvested from mice. (D) Relative expression of miR-1a in ventricles of mice subjected to control or MI surgery. Data are expressed as means ± SEM; n=4 mice per group. \*\**P* < 0.01 vs. Control LNA-NC, %*P* < 0.05 vs. MI LNA-NC. Two-tailed unpaired t-test. (E) Relative expression of miR-15b in ventricles of mice subjected to control or MI surgery. Data are expressed as means ± SEM; n=4 mice per group; \*\**P* < 0.01 vs. Control LNA-NC, %*P* < 0.05 vs. MI LNA-NC. Two-tailed unpaired t-test. (F) Longitudinal monitoring of cardiac % Ejection fraction (%EF). Data are expressed as means ± SEM; N=4/5 mice per group; \*\**P* < 0.01 vs. MI LNA-NC. Two-tailed unpaired t-test. (G) Survival curve in MI group. N=4/5 mice per group. (H) The mouse heart was divided into the Remote and Border zone, as depicted. (I) Heat map shows the mRNA expression levels of pathological remodeling marker genes (*Nppa*, *Myh7* and *Myh7/Myh6 ratio*) and fibrotic remodeling marker genes (*Tgfβ2*, *Col3a1* and *Thbs1*) after LNA-NC or LNA-1a/15b injection and MI injury in the Remote and Border zone. Data was normalized to Control LNA-NC. The red and green colors indicate high and low expression values, respectively. n=4 mice per group. Two-tailed unpaired t-test. (J) Representative hematoxylin and eosin (HE) staining and picrosirius red stained heart sections at 28-days post MI surgery. (K) Quantitative cardiac fibrosis analysis. Data are expressed as means ± SEM; n=4 mice per group; \*\**P* < 0.01 vs. Control LNA-NC, %*P* < 0.05 vs. MI LNA-NC. Two-tailed unpaired t-test. MI, Myocardial Infarction.

Maintenance of cardiac function and the absence of mortality in MI mice treated with LNA-1a/15b indicates a possible attenuation of cardiac fibrosis and stiffness in these mice. Thus, we profiled key fibrotic marker genes including *transforming growth factor β2* (*Tgfβ2*), *Collagen type III alpha 1 chain* (*Col3a1*) and *thrombospondin 1* (*Thbs1*). As noted in **Figure 2I**, fibrotic marker gene expression was significantly reduced in MI mice treated with LNA-1a/15b compared to LNA-NC controls. Consistent with the negligible impact of the infarct on the remote zone, fibrotic marker gene expression in the remote zone was indistinguishable between control and MI mice, regardless of the treatment regime (**Figure 2I**). Finally, we stained sections of the respective hearts with Picrosirius Red, an established marker for tissue fibrosis ^31^. As noted in **Figure 2J, K**, LNA-1a/15b treated MI mice exhibited considerably less cardiac fibrosis compared to LNA-NC treated controls. Consistent with this observation, staining for the apoptosis marker cleaved-Caspase 3, revealed increased cell death in mouse LNA-NC treated hearts subjected to MI, but not in hearts treated with LNA-1a/15b (**Figure S6**). In sum, LNA-1a/15b treatment significantly attenuated the development of cardiac fibrosis, and its progression to heart failure and death in a mouse model of STEMI.

### Combinatorial miR-1a/15b interference specifically drives cardiomyocyte proliferation *in vivo* with minimal off-target effects

To ascertain if the beneficial effects of combinatorial miR-1a and miR-15b depletion is linked to adult cardiomyocyte proliferation, we examined distribution of the mitotic S-phase marker Ki67 and the metaphase marker phosphorylated Histone H3 (pHH3) in heart biopsies of the respective groups. Increased Ki67 and pHH3 signal was detected in cardiomyocytes of both sham and MI LNA-1a/15b treated mice (**Figure 3A-D; Figure S7A, B**), with greater levels of Ki67 and pHH3 induction observed in MI operated mice. Strikingly, when the respective biopsies were assessed for expression of the cytokinesis marker Aurora B, as readout for cell-cycle completion and daughter cell formation, we only detected a significant increase in the fraction of Aurora B^+^ cardiomyocytes in hearts of mice subjected to MI and treated with LNA-1a/15b (**Figure 3B, D**).

**Figure 3.**
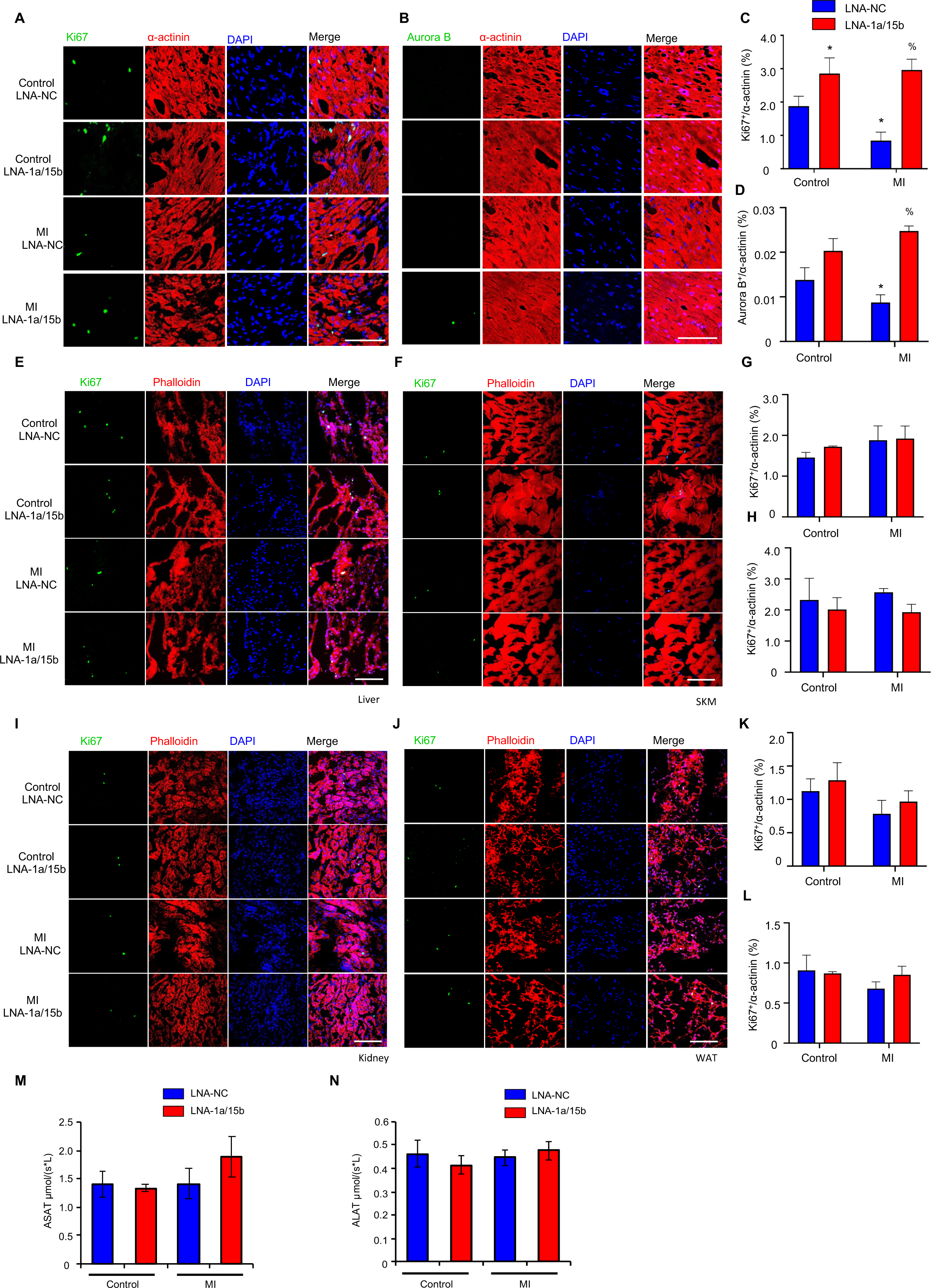
The effects of combinatorial miR-1a/15b interference on proliferation in cardiac tissue and in non-cardiac tissues *in vivo*. (A) Representative immunofluorescence images of Ki67 on heart sections of adult hearts injected with LNA-1a/15b or control 28-days post MI injury. Ki67 labels proliferating cells (green); cardiomyocyte-specific α-actinin (red) and DAPI (Blue). (B) Representative immunofluorescence images of Aurora B on heart sections of adult hearts injected with LNA-1a/15b or control 28-days post MI injury. Aurora B labels proliferating cells (green); cardiomyocyte-specific α-actinin (red) and DAPI (Blue). (C) Quantification of percentages of Ki67^+^ cardiomyocytes. Data are expressed as means ± SEM; N=3 mice per Control group, n=5 mice per MI group; \**P* < 0.05 vs. Control LNA-NC, %*P* < 0.05 vs. MI LNA-NC. Two-tailed unpaired t-test. Scale bar is 100 µm. (D) Quantification of percentages of Aurora B^+^ cardiomyocytes. Data are expressed as means ± SEM; N=3 mice per Control group, N=5 mice per MI group; %*P* < 0.05 vs. MI LNA-NC. Two-tailed unpaired t-test. Scale bar is 100 µm. (E) Representative images of Ki67 (green), Phalloidin (red) and DAPI (blue) in liver. Scale bar is 100 µm. (F) Representative images of Ki67 (green), Phalloidin (red) and DAPI (blue) in SKM. Scale bar is 100 µm. (G) Quantification of percentages of Ki67^+^ liver cells. Data are expressed as means ± SEM; n=3 mice per group. Two-tailed unpaired t-test. (H) Quantification of percentages of Ki67^+^ SKM cells. Data are expressed as means ± SEM; n=3 mice per group. Two-tailed unpaired t-test. (I) Representative images of Ki67 (green), Phalloidin (red) and DAPI (blue) in kidney. Scale bar is 100 µm. (J) Representative images of Ki67 (green), Phalloidin (red) and DAPI (blue) in WAT. Scale bar is 100 µm. (K) Quantification of percentages of Ki67^+^ kidney cells. Data are expressed as means ± SEM; n=3 mice per group. Two-tailed unpaired t-test. (L) Quantification of percentages of Ki67^+^ WAT cells. Data are expressed as means ± SEM; n=3 mice per group. Two-tailed unpaired t-test. SKM, Skeletal Muscle; WAT, White Adipose Tissue. (M) ASAT and (N) ALAT levels in plasma of C57BL/6J mice injected with wither LNA-1a/15b or Control LNA-NC 28-days post MI injury. Data are expressed as means ± SEM; n=5-7 mice per group. MI, Myocardial Infarction; ASAT, Aspartate aminotransferase; ALAT, Alanine aminotransferase.

LNAs are delivered systemically and can detrimentally affect non-target tissues. This is particularly relevant in regenerative- or proliferative-type therapies where proliferation activators targeting specific cell-types, may in parallel induce neoplasms formation due to mitotic activation in non-target cells. Thus, a degree of cell-type specificality in therapeutic response would be prudent. In **Figure S4A, B**, we demonstrated a lack of LNA-1a/15b impact on cardiac fibroblasts proliferation. To extend these findings, we asked how LNA-1a/15b inactivation would impact tissue proliferation of key non-cardiac cell-types in mice. As noted in **Figure 3E-L** and **Figure S8A-H**, negligible changes in cell proliferation, as determined by Ki67 and pHH3 staining, was detected in tissues including liver, skeletal muscle (SKM), kidney and white adipose tissue (WAT) of mice subjected to sham or MI surgery, in the absence or presence of LNA-1a/15b treatment.

Changes in cell and tissue metabolism is another important feature of neoplastic transformation and is broadly characterized by a shift from glucose and fatty acid oxidation as the primary mode of ATP generation to glycolysis, a process less efficient in ATP generation but advantageous for the generation of nucleotides, amino acids and phospholipids necessary building-blocks of cell proliferation ^32,33^. Given this, we performed metabolomic analysis on non-cardiac tissue namely the liver, lung, kidney, spleen and white adipose tissue (WAT) **(Figure S9A-E)**. Principal Component Analysis (PCA) revealed broad and overlapping signals distribution across the respective groups in liver, lung, kidney and spleen biopsies, suggestive of insignificant deviation from Control LNA-NC treated mice. In contrast, analysis of WAT biopsies from the respective groups revealed unique signal clustering of biopsies derived from LNA-NC mice subjected to MI compared to the other groups. Importantly, WAT biopsies harvested from mice subjected to MI and treated with LNA-1a/15b exhibited pronounced signal overlap with Control LNA-NC treated tissue **(Figure S9A-E)**. Taken together, combinatorial inhibition of miR-1a and miR-15b caused negligible off-target effects on non-cardiac tissue metabolism and proliferation rates.

Finally, to understand any possible effect of LNA-1a/15b therapy on systemic toxicity, we assessed circulating levels of biomarker enzymes Aspartate aminotransferase (ASAT) and Alanine aminotransferase (ALAT) ^75^. While ASAT is commonly associated with liver and heart damage, it can also be found in other tissues, including skeletal muscle and kidneys. ALAT is primarily found in hepatocytes. Upon tissue damage due to drug toxicity, these enzymes are released into the blood. Upon tissue damage due to drug toxicity, release these enzymes into the blood. Thus, circulating ASAT and ALAT levels in the blood directly correlates with tissue damage and toxicity ^76^. As noted in **Fig. 3M, N**, LNA-1a/15b had minimal impact on circulating ASAT and ALAT levels compared to controls, indicative of an acceptable safety profile. In sum, these data indicate that LNA-1a/15b treatment is non-toxic and provokes minimal tissue off-target effects.

### Combinatorial miR-1a/15b interference enhances cardiomyocyte proliferation and function in human cardiac tissue mimetics

In order to understand the potential of LNA-1a/15b therapy for clinical translation, we first assessed the effects of LNA-1a/15b in the human induced pluripotent stem cells-derived cardiomyocytes (hiPSC-CMs) cultured under control normoxia or in hypoxia (1%O2) as an *in vitro* model for myocardial infarction ^22^. LNA-1a/15b treatment effectively suppressed RNA levels of miR-1a and miR-15b in the hiPSC-CMs (**Figure S10A, B**) in normoxia and hypoxia. As observed in **Fig. S10C, D**, LNA-1a/15b treatment led to increased Aurora B signal, indicative of a conserved function for LNA-1a/15b in driving mature human cardiomyocyte proliferation and cell-cycle completion. Next, we utilized human iPSC-derived 3D cardiac tissue mimetics to determine the proliferative impact of LNA-1a/15b treatment in tissue. Utilizing a variation of a previously established 3D cardiac mimetic model with inclusion of a maturation phase ^34–36^, we were able to generate adult cardiac mimetics exhibiting characteristics and gene expression profiles characteristic of adult tissue ^37^ (**Figure S11A-H**). In this setting, cardiac mimetics were cultured under control normoxia or in hypoxia, treated with LNA-NC or LNA-1a/15b and assessed for proliferation by EdU-incorporation (as readout for S-Phase entry and DNA replication) and pHH3 staining (**Figure 4A-D)**. As observed in the mouse STEMI model, a similar pattern of proliferative induction by LNA-1a/15b was detected in both control normoxia and hypoxic conditions, with elevated levels of proliferation observed in hypoxia.

**Figure 4.**
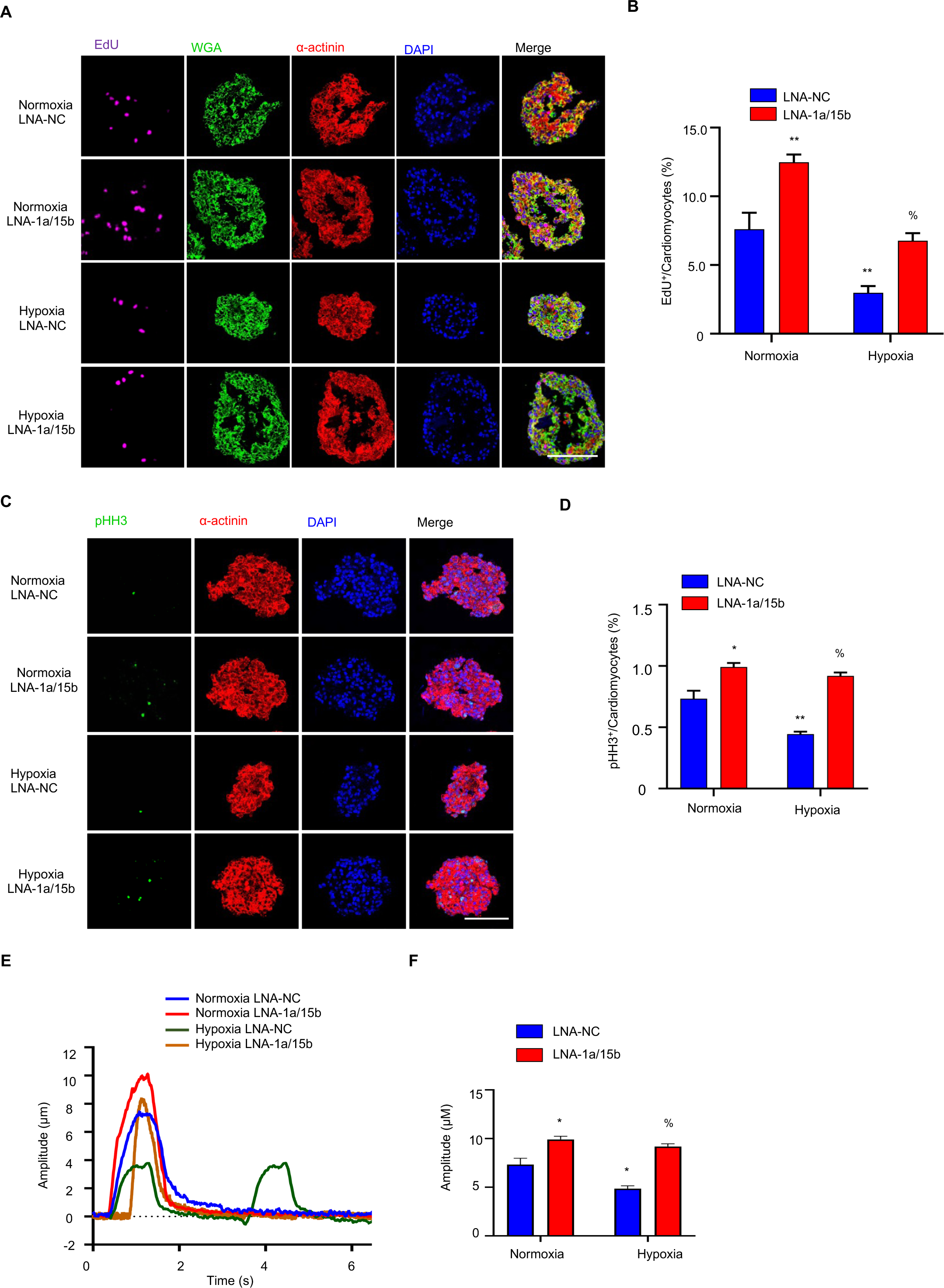
The effects of combinatorial miR-1a/15b interference on cardiomyocyte proliferation and cardiac function in human cardiac tissue mimetics. (A) Representative immunofluorescence images of EdU incorporation on sections of 40-day-old **human cardiac mimetics** treated with LNA-1a/15b or LNA-NC in both normoxia and hypoxia. EdU labels proliferating cells (Magenta); Wheat-Germ-Agglutinin (WGA) marks cell surfaces (green); cardiomyocyte-specific α-actinin (red) and DAPI (Blue). (B) Quantification of percentages of EdU^+^ cardiomyocytes. Data are expressed as means ± SEM; n=3 per group; \*\**P* < 0.01 vs. Normoxia LNA-NC, %*P* < 0.05 vs. Hypoxia LNA-NC. Two-tailed unpaired t-test. Scale bar is 100 µm. (C) Representative immunofluorescence images of pHH3 on sections of 40-day-old human cardiac mimetics treated with LNA-1a/15b or LNA-NC in both normoxia and hypoxia. pHH3 labels proliferating cells (Magenta); cardiomyocyte-specific α-actinin (red) and DAPI (Blue). Scale bar is 100 µm. (D) Quantification of percentages of pHH3^+^ cardiomyocytes. Data are expressed as means ± SEM; n=3 per group; \*\**P* < 0.01 vs. Normoxia LNA-NC, %*P* < 0.05 vs. Hypoxia LNA-NC. Two-tailed unpaired t-test. (E) Representative traces of contractile human cardiac mimetics. Human cardiac mimetics were treated with LNA-1a/15b or LNA-NC for 3 days in both normoxia and hypoxia, and then the contractility assays were performed by determining the amplitude peak of contracting human cardiac mimetics. (F) Quantification of the amplitude peak of contracting human cardiac mimetics. Data are expressed as means ± SEM; 4 - 5 organoids per group; \**P* < 0.05 vs. Normoxia LNA-NC, %*P* < 0.05 vs. Hypoxia LNA-NC. Two-tailed unpaired t-test.

Furthermore, as observed in the mouse studies, LNA-1a/15b treatment of human organoids subjected to severe hypoxic stress prevented the onset of contractile dysfunction (**Figure 4E, F).** Calcium flux measurements were performed to follow to the contractility patten of organoids from the respective groups (**Fig. 4E**), and quantified as shown in **Fig. 4F**. Cardiac organoids cultured in hypoxia revealed the expected decrease in contractile amplitude concomitant to increased cycling frequency (**Fig. 4E, F**). However, LNA-1a/15b treatment in human cardiac mimetics rescued the loss in contractility induced by severe hypoxic stimulation (**Fig. 4E, F**). Consistent with previous data, LNA-1a/15b treatment effectively suppressed RNA levels of miR-1a and miR-15b in the human cardiac mimetics (**Figure S12A, B**). In sum, our data indicates that LNA-1a/15b drives mitotic entry and cell-cycle completion in to maintain cardiac function in mature human tissue in the face of hypoxic insult.

### Mechanistic basis of LNA-1a/15b function in cardiomyocyte proliferative induction

Whilst the vast majority of therapeutic interventions involving miRNA inhibition rely on targeted inactivation of a single miRNA, our screening efforts reveals that combinatorial approaches can potentially provide greater efficacy and robustness in driving adult cardiomyocyte proliferation. Further, this combinatorial effect is not restricted to rodents, rather it is conserved across species as noted by the effects of miR-1a/15b inactivation in mouse, rat and human cardiomyocytes and cardiac tissue (**Fig. 1D-G; Fig. S5E, F; Fig. 3A-D; Fig. S7A, B; Fig. 4A-D**). Strikingly, due to the enrichment of miR-1a and miR-15b in cardiomyocytes, we detected negligible effect of LNA-1a/15b treatment on proliferative induction of fibroblasts, and on the heterogenous population of cells contained within non-cardiac tissue (**Fig. S4A, B; Fig. 3E-L; Fig. S8A-H**). The specificity of LNA-1a/15b function is also reflected at the level of the respective non-cardiac tissue metabolomes (**Fig. S9A-E**). Given this, we sought to understand the downstream consequence of miR-1a/15b inhibition on coding gene regulators, as mediators of proliferative induction, specifically in cardiomyocytes. Thus, post-mitotic P7 cardiomyocytes were treated with LNA-NC, LNA-1a, LNA-15b or LNA-1a/15b and hypoxia stimulated (to mimic the MI setting). As shown in the heatmap on **Fig. 5A**, differential gene expression analysis revealed specific changes in gene expression unique to cardiomyocytes stimulated with the combination of LNAs targeting both miR1a/15b, but not in control LNA-NC, or in cardiomyocytes treated with LNAs targeting only miR-1a or miR-15b individually. Next, Kyoto Encyclopedia of Genes and Genomes (KEGG) clustering analysis revealed enrichment of genes linked to cardiac development, cell/tissue homeostasis, and cardiac growth and morphogenesis (**Fig. 5B; Fig. S13A-G**). Consistent with this data, Gene Set Enrichment Analysis (GSEA) enrichment analysis of all differentially expressed genes between LNA-NC, LNA-1a, LNA-15b and LNA-1a/15b treated groups revealed distribution into cardiac linked pathways (**Fig. 5C**). Given that the LNA-1a/15b combination proved most relevant by virtue of its ability to maintain cardiac function and organismal survival in the face of MI, we sub-classified genes within the respective Gene Ontology (GO) pathways into 4 groups comprising of genes regulated specifically by miR-1a or miR-15b depletion, respectively (miR-1a and miR-15b only), genes regulated in-common by both miR-1a and miR-15b depletion (Common), and differentially expressed genes specific to combinatorial inhibition of miR-1a and miR-15b (Unique) (**Fig. 5C**). As expected, regulation of the vast majority of genes can be linked directly to miR-1a and/or miR-15b function (**Fig. 5C; Fig. S14A-H**). Surprisingly, a fairly large number of differentially expressed genes could neither be linked to direct miR-1a or miR-15b function, based on known and predicted targets of the respective miRNAs and our expression data (**Fig. S14D, H**) ^21^. Genes depicted in the **Fig. S14D** heatmap encompass miR-1a/15b regulated genes that are unaffected or only mildly altered (and below the significance threshold) upon treatment with miR-1a LNAs or miR-15b LNAs individually. Many of the genes within this Unique subgroup are characterized by a role in inducing cardiomyocyte de-differentiation, early cardiac development and remodeling comprising of genes such as T-Box Transcription Factor 20 (TBX20) ^38,39^, PR-Domain Zinc Finger Protein 16 (PRDM16) ^40^, ADP-Ribosylhydrolase Like 1 (ADPRHL1) ^41^, Glycogen Synthase Kinase 3 Beta (GSK3B) ^42^, Fibroblast Growth Factor Receptor Substrate 2 (FRS2) ^43^, Sorbin And SH3 Domain Containing 2 (SORBS2) ^44^, WW Domain Containing Transcription Regulator 1 (WWTR1) ^45^ and SRY-Box Transcription Factor 9 (SOX9) ^46^ (**Fig. S14D**). Connected to these are modulators of sarcomerogenesis, cardiac calcium handling and contractility including Cysteine And Glycine Rich Protein 3 (CSRP3) ^47^, Synaptopodin 2 Like (SYNPO2L) ^48^, Myosin XVIIIB (MYO18B) ^49,50^, Xin Actin Binding Repeat Containing 1 (XIRP1) ^51^, NADPH Oxidase 4 (NOX4) ^52^, Calcium Voltage-Gated Channel Subunit Alpha1 C (CACNA1C) ^53^, Mitochondrial Dynamin Like GTPase (OPA1) ^54^, Ankyrin 2 (ANK2) ^55^, PDZ And LIM Domain 7 (PDRM17) ^56^ and Ryanodine Receptor 2 (RYR2) ^57^ (**Fig. S14D**). Further, we detected upregulation of drivers of angiogenesis, lymphangiogenesis, neurogenesis and extracellular matrix and stromal cell activators consisting of Prospero Homeobox 1 (PROX1) ^58^, Neuropilin 2 (NRP2) ^59^ and Transforming Growth Factor Beta 3 (TGFB3) ^60^ (**Fig. S14D**). Finally, this Unique group contained general transcriptional and translational modulators, and epigenetic regulators including key factors such as AT-Rich Interaction Domain 1A (ARID1A) ^61^, Polybromo 1 (PBRM1) ^62^, Mediator Complex Subunit 1 (MED1) ^63^, Lysine Demethylase 6B (KDM6B) ^64^, and lesser understood components including Zinc Finger MIZ-Type Containing 1 (ZMIZ1) ^65^ and Dishevelled Segment Polarity Protein 3 (DVL3) ^66^ (**Fig. S14D**). Taken together, these data suggest a possible molecular synergism wherein simultaneous depletion of miR-1a and miR-15b facilitates the specific activation of genes, which in this context serves to provide a pro-proliferative environment in post-mitotic adult cardiomyocytes by co-opting key accessory pathways driving adult cardiomyocyte de-differentiation and plasticity, remodeling of the cardiomyocyte sarcomere and contractile machinery, concomitant to the induction of cardiac angiogenesis, neurogenesis and stromal activation to support survival of new cardiomyocytes, to enable effective adult cardiac regeneration.

**Figure 5.**
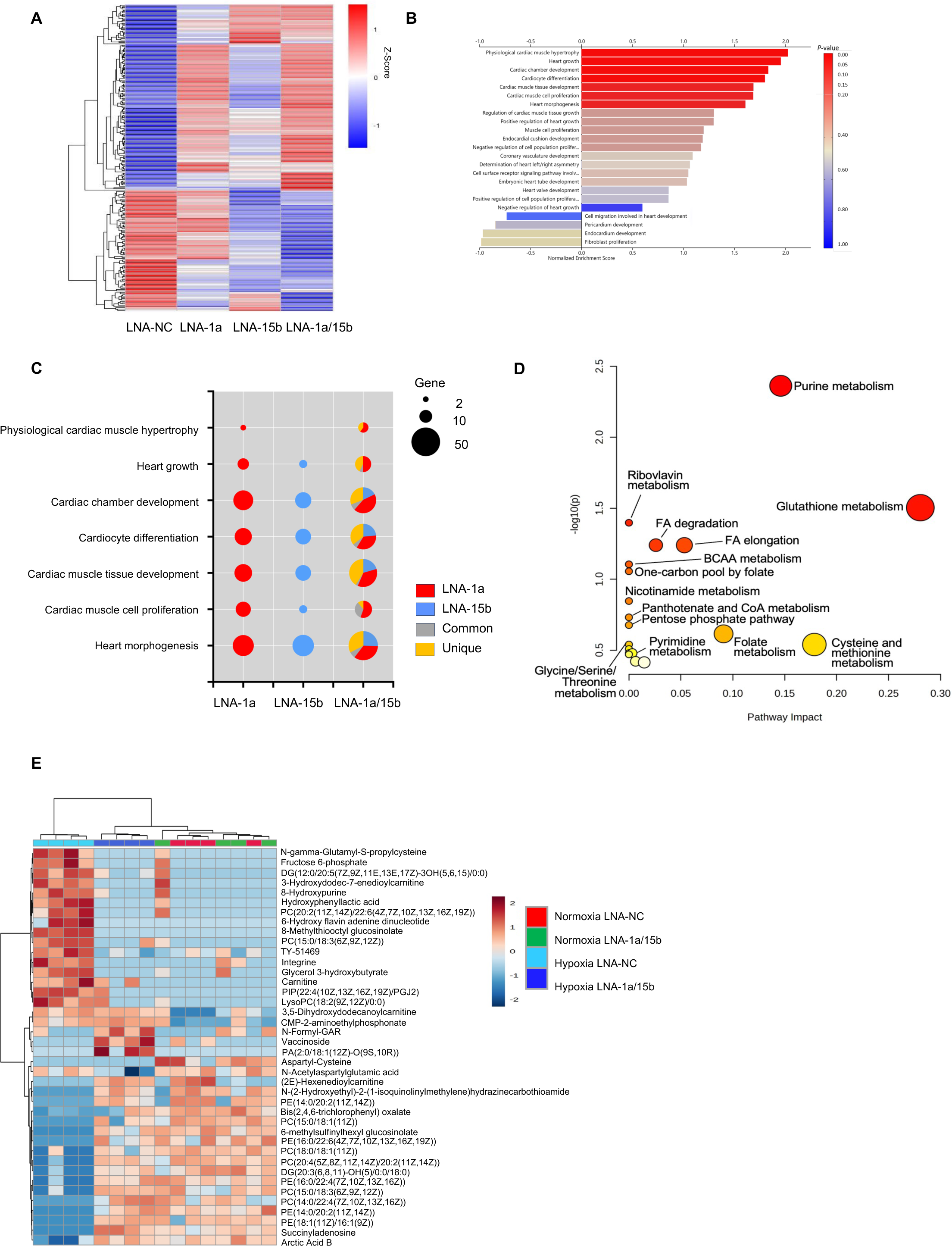
Changes in gene expression and metabolites with inhibition of miR-1a and miR-15b. (A) The heatmap shows the variant genes. X-axis: samples of LNA-NC, LNA-1a, LNA-15b, LNA-1a/15b; The red and blue colors indicate upregulation and downregulation, respectively. Means of n = 3 biological replicates per group. (B) Gene Ontology (GO) biological process terms enriched in LNA-1a/15b-regulated genes. (C) Top 7 GO biological process terms related to the heart development enriched in LNA-1a, LNA-15b and LNA-1a/15b. Red, genes regulated specifically by miR-1a inhibition; Blue, genes regulated specifically by miR-15b inhibition; grey, genes regulated in-common by both miR-1a and miR-15b depletion; yellow, genes regulated unique to combinatorial inhibition of miR-1a and miR-15b. (D) Pathway analysis bubble plot in KEGG shows the enriched metabolic pathways in P1 rat cardiomyocytes treated with LNA-1a/15b under hypoxia. The horizontal axis is the pathway impact, which represents the importance of differential metabolites in metabolic pathways. The vertical axis represents the results of pathway enrichment analysis. The dot size represents the centrality of the metabolites involved in corresponding metabolic pathway. The color of the dot represents the impact factor; large sizes and dark colors represent the central metabolic pathway enrichment and pathway impact values, respectively. (E) Heat map of relative metabolite abundance in Neonatal P1 rat cardiomyocytes treated with either control LNA-NC or LNA-1a/15b, and then incubated in the hypoxia and hypoxia for 2 days. Depicted are metabolites with log2(fold change)>0.5 compared to Normoxia LNA-NC treatment and adjusted p value<0.01 in at least one treatment group compared to corresponding control LNA-NC. n=4 biological replicates per group.

Given the crucial role of the cell metabolome in enabling and facilitating cell proliferation ^67^, we analyzed changes in cellular metabolite composition as a result of LNA-1a/15b treatment under basal and hypoxic stress conditions. As noted in **Fig. 5D, E**, depletion of miR-1a/15b in hypoxic cardiomyocytes led to elevation of metabolites necessary for proliferative induction and cell-cycle completion, including metabolites of purine and pyrimidine biosynthesis ^68,69,70,71^, branched chain amino acid (BCAA) metabolites ^32,70^, nicotinamide and riboflavin components ^32,70^, panthotenate and co-enzyme A metabolites necessary for acylation of metabolic intermediates ^68,71^, folate and 1-carbon pathway metabolites ^70,71^, phosphatidylcholines ^72^ and glycerol hydroxybutyrates ^32,73,74^. Strikingly, there was little evidence for elevation of these metabolites neither in response to hypoxia stimulation alone nor upon LNA-NC treatment (**Fig. 5D, E**). Thus, these data suggest that LNA-1a/15b serves to very specifically reprogram the cardiac transcriptome and metabolome to enable and support cardiac tissue regeneration.

## Discussion

miRNAs are known to play critical roles in cardiac pathophysiology, including MI and heart failure. Most studies have primarily focused on the role of individual miRNAs, although combinatorial activities of miRNAs are being increasingly investigated and recognized in recent years ^77,78^. Although the impact of individual miRNA modulation cannot be underestimated, we speculate that more complex processes might necessitate the simultaneous combinatorial modulation of two or more regulatory moieties. Given the complexity and robustness engrained within adult cardiomyocytes to maintain terminal differentiation, structural integrity within the tissue complex and interaction with surrounding cardiomyocytes, stromal and mural cells in order to facilitate cardiac contractility and function, our observations indicate that the induction and re-entry of post-mitotic adult cardiomyocytes could be one such example to benefit from complex and combinatorial miRNA modulation. This is supported by recent data where combinatorial modulation of coding genes through simultaneous co-expression of four genes (comprising transcription factors and cell-cycle regulator combinations) have been shown necessary to drive adult cardiomyocyte mitotic re-entry ^79–81^. Although these finding are of key importance in understanding mechanisms underlying induction of adult cardiomyocyte mitosis, translating these findings into a clinically feasible strategy is challenging due to the need for cardiac directed gene therapy (necessitating the need for invasive delivery procedures) and the clear risk of therapy leakiness which could lead to neoplasm formation in non-cardiac tissue (due to the ubiquitous nature and function of the transcription factors and cell-cycle regulator combinations ectopically expressed in these strategies ^79–81)^. Thus, developing clinically feasible and accessible therapeutic strategies for cardiac regeneration remains an unmet need. Combinatorial miRNAs therapy targeting cardiac enriched miRNAs may offer such an avenue by virtue of the ability to inactivate these miRNAs through systemic delivery of stabilized anti-sense moieties, thus removing the need for invasive procedures such as catheter-based delivery or direct cardiac injection ^82,83^.

According to the Human microRNA Disease Database (HMDD) microRNA–disease associations, the 17 miRNAs we selected are upregulated in the patients with MI and heart failure ^21^. This is consistent with our data demonstrating elevation of all 17 miRNAs heart biopsies of patients with myocardial infarction (**Figure S1A**). Among the proliferation-regulating miRNAs, we have used a combinatory approach to identify a combination that could further increase cardiomyocyte proliferation compared to individual miRNA. Our study identified that simultaneous inhibition of miR-1a and miR-15b increased cardiomyocyte proliferation and improved cardiac function *in vitro* and *in vivo*. Both of miR-1a and miR-15b are highly expressed in hearts, and they have been documented to regulate cardiomyocyte proliferation ^84,85^. Overexpression of miR-1 in the developing hearts leads to decreased cardiomyocyte proliferation ^86,87^.

Conversely, miR-1a deletion increases cardiomyocyte proliferation in the adult hearts ^85^, and attenuates cardiac I/R injury in mice and rats ^88,89^. miR-15b is upregulated in the infarcted zone of porcine cardiac tissue in response to ischemic injury, and miR-15b inhibition increases cardiac regeneration and protects against ischemic-induced injury ^84^. Our data showed that combinatory inhibition of miR-1a and miR-15b could further increase cardiomyocyte proliferation in P1 and P7 rat cardiomyocytes, and could enhance cardiomyocyte proliferation and improve cardiac function in mice *in vivo*. As access to cardiac tissue, in particular, is challenging due to ethical concerns, human cardiac organoids serve as proxy material in which functionality of this therapeutic approach can be assessed. In utilizing iPSC-derived human cardiac mimics, we were able to demonstrate a conservation of function of inhibition of miR-1a and miR-15b in driving adult human cardiomyocyte proliferation, leading to maintenance of cardiac contractility in the face of hypoxic insult. Therefore, combinatorial miRNAs therapy may provide therapeutic benefit by simultaneously interfering with the many dysregulated pathways in myocardial infarction and heart failure. In sum, the present finding: (i) LNA-1a/15b synergistically enhanced cardiomyocyte proliferation *in vitro* and *in vivo*; and (ii) LNA-1a/15b synergistically reduced MI-induced EF and fibrosis, and improved cardiac function in mice *in vivo*; and (iii) LNA-1a/15b synergistically increased cardiomyocyte proliferation and improved contractility in human cardiac mimetics. Our study thus reveals a novel mode of two-miRNA combination that offers synergistic regulation on cardiac regeneration and enhanced cardiac function in mice following MI.

miR-1 and miR-15b are expressed in a highly tissue-restricted manner. miR-1 and miR-15b are predominantly expressed in mouse and human heart ventricle, with low levels detected in other tissues ^90,91^. In the present study, the use LNA-modified anti-miRs in targeting miR-1a and miR-15b, facilitates indirect tissue-targeting due the predominant expression of miR-1a and miR-15b in heart tissue. In accord, macroscopic analysis of morphology and structure of non-cardiac tissue, including the lung, spleen, SKM, liver and WAT revealed negligible differences between LNA-1a/15b and LNA-NC treated mice. Taken together, the synergism offered through simultaneous inhibition miR-1a and miR-15b, combined with the restricted expression of miR-1a and miR-15b, may facilitate tissue-specific precision therapy, to lower side effects and the possibility of neoplasm formation in other tissues. Thus, inhibiting miR-1a and miR-15b is a promising strategy to enhance cardiac regeneration in patients of heart failure.

Currently, the LNA-based anti-miRs have been widely used to suppress the function of miRNAs, and have also been shown to be safe and effective in human studies ^92–94^. Various studies have been convincingly shown that targeting miRNAs may improve cell therapy or enhance endogenous cardiac repair processes, and LNA-based anti-miRs have already been used in large animal models and a first clinical trial. For example, LNA-anti-miR-21 treatment reduces cardiac fibrosis and hypotrophy and improves cardiac function in a pig model of Ischemia/reperfusion (I/R) injury ^95^. In addition, LNA-based anti-miR-132 treatment improves EF and ameliorates cardiac dysfunction in response to MI in a porcine model ^92^. Furthermore, a recent clinical trial involving delivery of a synthetic LNA-based antisense oligonucleotide against miR-132 (anti-miR-132) in heart failure patients (NCT04045405) revealed a potential benefit of CDR132L in attenuating heart failure ^92^. MRG-110, an LNA-based antisense oligonucleotide targeting miR-92a, has been also tested in healthy human adults and MRG-110 treatment reduces miR-92a level and de-represses the target genes in human peripheral blood cells ^93^. These studies indicate that further development of LNA-based anti-miR therapies could provide a route to developing efficacious therapies, of which LNA-anti-miR-1a/15b would be one such strategy.

Taken together, combinatorial LNA-anti-miR-1a/15b treatment results in pro-proliferative effects on adult cardiomyocytes and improved cardiac function in response to myocardial infarction. Our data provides a novel and clinically feasible LNA-based anti-miR-1a/15b strategy for treatment of STEMI, and potentially heart failure through re-activation of adult cardiomyocyte mitotic re-entry and cardiac regeneration.

## Data Availability

All data produced in the present study are available upon reasonable request to the authors.

## Acknowledgements

We thank P. Schuster, H. Fischer, all members of the Institute for Cardiovascular Regeneration and Genome Biologics for their technical, regulatory, administrative, analytical, reagent and scientific support. We are grateful to A. Fischer for performing the surgeries. This work was supported by the Messer Foundation, LOEWE Center for Cell and Gene Therapy and the European Innovation Council (GA: 822455) to J.K., and the SFB-TRR 267 (Non-coding RNA in the cardiovascular system) and the German Research Foundation (DFG) (Exc2026) to S.D. and J.K., and instrument grant support (INST 515/28-1 FUGG) to M.P., the European Research Council (Angiolnc) to S.D., and the Foundation for Pathobiochemistry and Molecular Diagnostics to P.M. Y.W. was supported by the China Scholarship Council (CSC) Grant #202108080020.

## Author Contributions

T.Y., C.Z., P.M.D., K.B., Y.M., A.P.D. and P.M. conducted experiments and analyzed results. H.Y. performed the bioinformatics analyses. Experiments were designed by A.Z., S.D. and J.K.. T.Y. and J.K. wrote the manuscript with support from J.W..

## Supplementary Figure Legends

**Supplementary Figure S1. Expression levels of the miRNAs**

(A) Relative expression of miRNAs in heart biopsies from human patients with MI and healthy controls. Data are expressed as means ± SEM; n=4 for healthy controls, n≥6 for MI group; \*\**P* < 0.01, \**P* < 0.05 vs. healthy heart. All by Mann Whitney test. (B) Relative miRNA expression levels were quantified by qRT-PCR in the neonatal P1 rat cardiomyocytes after 48 h treatment with pool of 17 LNAs. Data are expressed as means ± SEM; n=3; \**P*<0.05, \*\**P*<0.01 vs. LNA-NC. Two-tailed unpaired t-test.

**Supplementary Figure S2. Effect of inhibition of miRNAs in the rat cardiomyocytes**

(A) Immunofluorescence images of Aurora B localization at the cleavage furrow at the telophase during mitosis. Aurora B (green) and α-actinin (red), and DAPI (blue). Scale bar is 100 µm. (B) Quantification of the percentage of Aurora B^+^ cells in α-actinin+ cardiomyocytes for the combination LNA-1a/15b. Data are expressed as means ± SEM; n=3; \**P*<0.05 vs. LNA-NC. Two-tailed unpaired t-test. (C) Quantification of the percentage of Aurora B^+^ cells in α-actinin+ cardiomyocytes for the combination LNA-1a/15b/15b. Data are expressed as means ± SEM; n=3; \**P*<0.05 vs. LNA-NC. Two-tailed unpaired t-test. (D) Quantification of the percentage of Aurora B^+^ cells in α-actinin+ cardiomyocytes for the combination LNA-1a/15b/15b. Data are expressed as means ± SEM; n=3. \**P*<0.05 vs. LNA-NC. Two-tailed unpaired t-test. (E) Relative miRNA expression levels were quantified by qRT-PCR in the P1 rat cardiomyocytes after 48 h treatment with combination LNA-1a/15b. Data are expressed as means ± SEM; n=3; \*\**P*<0.01 vs. LNA-NC. Two-tailed unpaired t-test. (F) Relative miRNA expression were levels quantified by qRT-PCR in the P1 rat cardiomyocytes after 48 h treatment with combination LNA-1a/15b/15b. Data are expressed as means ± SEM; n=3; \*\**P*<0.01 vs. LNA-NC. Two-tailed unpaired t-test. (G) Relative miRNA expression levels were quantified by qRT-PCR in the P1 rat cardiomyocytes after 48 h treatment with combination LNA-1a/15b/15b. Data are expressed as means ± SEM; n=3; \*\**P*<0.01 vs. LNA-NC. Two-tailed unpaired t-test.

**Supplementary Figure S3. miRNA expression levels in the P7 rat cardiomyocytes**

Relative miRNA expression levels were quantified by real-time PCR in the P7 rat cardiomyocytes after 48 h treatment with LNA-1a/15b. Data are expressed as means ± SEM; n=3; \*\**P*<0.01 vs. LNA-NC. Two-tailed unpaired t-test.

**Supplementary Figure S4. The effects of combination LNA-1a/15b in rat fibroblasts**

(A) Representative immunofluorescence images of EdU incorporation on rat fibroblasts treated with LNA-1a/15b or LNA-NC. EdU labels proliferating cells (Magenta); α-SMA marks fibroblast (green) and DAPI (Blue). Scale bar is 100 µm. (B) Quantification of percentages of EdU^+^ cardiomyocytes. Data are expressed as means ± SEM; n=3 per group. Two-tailed unpaired t-test.

**Supplementary Figure S5. Inhibition of miR-1a and/or miR-15b by LNA-modified oligonucleotides in mice *in vivo***

(A) Schematic representation of the experimental timeline of mice injected with control LNA-NC, LNA-1a, LNA-15b or LNA-1a/15b. The mouse hearts were harvested at day 7 post LNAs injection. (B-D) Relative miRNA expression levels were quantified by real-time PCR in the heart after 7 days injection with LNA-NC, LNA-1a, LNA-15b or LNA-1a/15b. Data are expressed as means ± SEM; n=3 mice per group; \*\**P*<0.01 vs. LNA-NC. Two-tailed unpaired t-test. (E) Representative immunofluorescence images of pHH3 on heart sections of adult hearts injected with LNA-1a, LNA-15b, LNA-1a/15b or control LNA-NC for 7 days. pHH3 labels proliferating cells (green); cardiomyocyte-specific α-actinin (red) and DAPI (Blue). (F) Quantification of percentages of pHH3+ cardiomyocytes. Data are represented as Mean±SEM; n=3 mice per group; \**P*< 0.05. vs. LNA-NC. Two-tailed unpaired t-test.

**Supplementary Figure S6. Detection of apoptotic cells by Cl. Caspase 3 and TUNEL staining**

Immunofluorescence images of Cl. Caspase 3 on heart sections from adult hearts injected with LNA-1a/15b or control LNA-NC 28-days post MI injury. DAPI labels nuclear (Blue); Tomato-lectin (Green) labels compact myocardium; Cl. Caspase 3 (Orange) label apoptotic cells. MI, Myocardial Infarction.

**Supplementary Figure S7. Inhibition of miR-1a and miR-15b by LNA-1a/15b in mice subjected to Myocardial Infarction**

(A) Representative immunofluorescence images of pHH3 on heart sections of adult hearts injected with LNA-1a/15b or control LNA-NC 28-days post MI injury. pHH3 labels proliferating cells (green); cardiomyocyte-specific α-actinin (red) and DAPI (Blue). Scale bar is 100 µm. (B) Quantification of percentages of pHH3^+^ cardiomyocytes. Data are expressed as means ± SEM; n=3 mice per Control group, n=5 mice per MI group; \**P* < 0.05 vs. Control LNA-NC, %*P* < 0.05 vs. MI LNA-NC. Two-tailed unpaired t-test. MI, Myocardial Infarction.

**Supplementary Figure S8. The effects of combinatorial miR-1a/15b interference on proliferation in non-cardiac tissues**

The liver, SKM, Kidney and WAT tissues from the LNA-NC or LNA-1a/15b treated mice were stained for pHH3 (green), Phalloidin (red) and DAPI (blue). (A) Representative images of pHH3 (green), Phalloidin (red) and DAPI (blue) in liver. Scale bar is 100 µm. (B) Representative images of pHH3 (green), Phalloidin (red) and DAPI (blue) in SKM. Scale bar is 100 µm. (C) Quantification of percentages of pHH3^+^ liver cells. Data are expressed as means ± SEM; n=3 mice per group. Two-tailed unpaired t-test. (D) Quantification of percentages of pHH3^+^ SKM cells. Data are expressed as means ± SEM; n=3 mice per group. Two-tailed unpaired t-test. (E) Representative images of Ki67 (green), Phalloidin (red) and DAPI (blue) in kidney. Scale bar is 100 µm. (F) Representative images of pHH3 (green), Phalloidin (red) and DAPI (blue) in WAT. Scale bar is 100 µm. (G) Quantification of percentages of pHH3^+^ kidney cells. Data are expressed as means ± SEM; n=3 mice per group. Two-tailed unpaired t-test. (H) Quantification of percentages of pHH3^+^ WAT cells. Data are expressed as means ± SEM; n=3 mice per group. Two-tailed unpaired t-test. SKM, Skeletal Muscle; WAT, White Adipose Tissue.

**Supplementary Figure S9. The effects of combinatorial miR-1a/15b interference on non-cardiac tissue metabolism**

Principal Component Analysis (PCA) of denoted cohorts on Liver (A), Lung (B), Kidney (C), spleen (D) and WAT (F). Control LNA-NC: n=4; Control LNA-1a/15b: n=4; MI LNA-NC: n=4-7; MI LNA-1a/15b: n=4-6. WAT, White Adipose tissue.

**Supplementary Figure S10. Inhibition of miR-1a and miR-15b by LNA-1a/15b in human iPSC-CMs**

(A) Relative miR-1 expression level was quantified by real-time PCR in the 40-day-old human iPSC-CMs 48h after LNA-1a/15b or LNA-NC treatment in both normoxia and hypoxia *in vitro*. Data are expressed as means ± SEM; n=3; \**P* < 0.05, \*\**P* < 0.01 vs. Normoxia LNA-NC, %*P* < 0.05 vs. hypoxia LNA-NC. Two-tailed unpaired t-test. (B) Relative miR-15b expression level was quantified by real-time PCR in the 40-day-old human iPSC-CMs 48h after LNA-1a/15b or LNA-NC treatment in both normoxia and hypoxia *in vitro*. Data are expressed as means ± SEM; n=3; \**P* < 0.05, \*\**P* < 0.01 vs. Normoxia LNA-NC, %*P* < 0.05 vs. hypoxia LNA-NC. Two-tailed unpaired t-test. (C) Representative immunofluorescence images of Aurora B (green), α-actinin (red) and DAPI in 40-day-old human iPSC-CMs 48h after LNA-1a/15b or LNA-NC treatment in both normoxia and hypoxia *in vitro*. (D) Quantification of percentages of Aurora B^+^ cardiomyocytes. Data are expressed as means ± SEM; n=3 per group; \**P* < 0.05 vs. Normoxia LNA-NC, %*P* < 0.05 vs. Hypoxia LNA-NC. Two-tailed unpaired t-test. Scale bar is 100 µm. Human iPSC-CMs, human-induced pluripotent stem cell-derived cardiomyocytes.

**Supplementary Figure S11. Maturation of human cardiac mimetics**

(A) Representative brightfield images of 40-day-old human cardiac mimeticss. Scale bar is 100 µm. (B) Representative immunofluorescence images of cardiomyocyte-specific α-actinin (green), DAPI (blue) and Phalloidin (Magenta) on whole human cardiac mimetics. Scale bar is 100 µm. (C-H) The human cardiac mimeticss were harvested at day 0 and day 40, and the gene expression levels including Myh6, Myh7, Mlc2a, Mlc2v, TTN-N2B, Cav3, NPPA, NPPB and PKM2for cardiomyocyte maturation were detected by the qRT-PCR. Data are expressed as means ± SEM; 50 organoids per group; \**P* < 0.05, \*\**P* < 0.01 vs. D0. Two-tailed unpaired t-test.

**Supplementary Figure S12. Inhibition of miR-1a and miR-15b by LNA-1a/15b in human cardiac mimetics**

(A) Relative miR-1 expression level was quantified by real-time PCR in the 40-day-old human human cardiac mimetics 96h after LNA-1a/15b or LNA-NC treatment in both normoxia and hypoxia *in vitro*. Data are expressed as means ± SEM; n=3; \**P*< 0.05, \*\**P* < 0.01 vs. Normoxia LNA-NC, %*P* < 0.05 vs. Hypoxia LNA-NC. Two-tailed unpaired t-test. (B) Relative miR-15b expression level was quantified by real-time PCR in the 40-day-old human human cardiac mimetics 96h after LNA-1a/15b or LNA-NC treatment in both normoxia and hypoxia *in vitro*.

**Supplementary Figure S13. Gene Set Enrichment Analysis (GSEA) enrichment plots related to the heart development**

(A) GSEA plot for GO class Physiological cardiac muscle hypertrophy in LNA-1a/15b group.. (B) GSEA plot for Heart growth in LNA-1a/15b group. (C) GSEA plot for Cardiac chamber development in LNA-1a/15b group. (D) GSEA plot for Cardiocyte differentiation in LNA-1a/15b group. (E) GSEA plot for Cardiac muscle tissue development in LNA-1a/15b group. (F) GSEA plot for Cardiac muscle cell proliferation in LNA-1a/15b group. (G) GSEA plot for Heart morphogenesis in LNA-1a/15b group.

**Supplementary Figure S14. Changes in the gene expression related to heart development with inhibition of miR-1a, miR-15b and miR-1a/15b**

(A-D) Heat map of RNA-seq expression z-scores computed for heart development related genes that are differentially expressed. Red and blue colors represent higher and lower relative expression levels, respectively. (A) Heat map shows that the genes are upregulated specifically by miR-1a depletion, but the genes are not significantly upregulated by miR-15b depletion. (B) Heat map shows that the genes are upregulated specifically by miR-15b depletion, but the genes are not significantly upregulated by miR-1a depletion. (C) Heat map shows that the genes are regulated in common by both miR-1a and miR-15b depletion. (D) Heat map shows that the genes are regulated unique to combinatorial inhibition of miR-1a and miR-15b, but the genes are unaffected or only mildly altered (and below the threshold of significance) upon miR-1a depletion or miR-15b depletion alone. Means of n = 3 biological replicates per group. (E-H) Enrichment analysis revealed the Gene Ontology (GO) biological process. (E) Top 10 GO-biological pathways of upregulated DEGs specifically in miR-1a depleted P7 rat cardiomyocytes. (F) Top 10 GO-biological pathways of upregulated DEGs specifically in miR-15a depleted P7 rat cardiomyocytes. (G) Top 10 GO-biological pathways of upregulated DEGs common to both miR-1a and miR-15b depleted P7 rat cardiomyocytes. (H) Top 10 GO-biological pathways of upregulated DEGs unique to combinatorial inhibition of miR-1a and miR-15b in P7 rat cardiomyocytes. DEGs, Differentially expressed genes.

## References

1. Vos T, Abajobir AA, Abate KH, Abbafati C, Abbas KM, Abd-Allah F, et al. Global, regional, and national incidence, prevalence, and years lived with disability for 328 diseases and injuries for 195 countries, 1990–2016: a systematic analysis for the Global Burden of Disease Study 2016. The Lancet 2017;390:1211–1259. doi:10.1016/S0140-6736(17)32154-2.

2. Mosterd A, Hoes AW. Clinical epidemiology of heart failure. Heart 2007;93:1137–1146. doi:10.1136/hrt.2003.025270.

3. Murry CE, Reinecke H, Pabon LM. Regeneration Gaps. Journal of the American College of Cardiology 2006;47:1777–1785. doi:10.1016/j.jacc.2006.02.002.

4. He L, Zhou B. Cardiomyocyte proliferation: remove brakes and push accelerators. Cell Res 2017;27:959–960. doi:10.1038/cr.2017.91.

5. Senyo SE, Steinhauser ML, Pizzimenti CL, Yang VK, Cai L, Wang M, et al. Mammalian heart renewal by pre-existing cardiomyocytes. Nature 2013;493:433–436. doi:10.1038/nature11682.

6. Bergmann O, Bhardwaj RD, Bernard S, Zdunek S, Barnabé-Heider F, Walsh S, et al. Evidence for Cardiomyocyte Renewal in Humans. Science 2009;324:98–102. doi:10.1126/science.1164680.

7. Bergmann O, Zdunek S, Felker A, Salehpour M, Alkass K, Bernard S, et al. Dynamics of Cell Generation and Turnover in the Human Heart. Cell 2015;161:1566–1575. doi:10.1016/j.cell.2015.05.026.

8. Ishikawa K, Weber T, Hajjar RJ. Human Cardiac Gene Therapy. Circ Res 2018;123:601– 613. doi:10.1161/CIRCRESAHA.118.311587.

9. High KA, Roncarolo MG. Gene Therapy. N Engl J Med 2019;381:455–464. doi:10.1056/NEJMra1706910.

10. Bischof C, Mirtschink P, Yuan T, Wu M, Zhu C, Kaur J, et al. Mitochondrial–cell cycle cross-talk drives endoreplication in heart disease. Sci Transl Med 2021;13:eabi7964. doi:10.1126/scitranslmed.abi7964.

11. Abplanalp WT, Fischer A, John D, Zeiher AM, Gosgnach W, Darville H, et al. Efficiency and Target Derepression of Anti-miR-92a: Results of a First in Human Study. Nucleic Acid Therapeutics 2020;30:335–345. doi:10.1089/nat.2020.0871.

12. Täubel J, Hauke W, Rump S, Viereck J, Batkai S, Poetzsch J, et al. Novel antisense therapy targeting microRNA-132 in patients with heart failure: results of a first-in-human Phase 1b randomized, double-blind, placebo-controlled study. European Heart Journal 2021;42:178–188. doi:10.1093/eurheartj/ehaa898.

13. Hinkel R, Batkai S, Bähr A, Bozoglu T, Straub S, Borchert T, et al. AntimiR-132 Attenuates Myocardial Hypertrophy in an Animal Model of Percutaneous Aortic Constriction. Journal of the American College of Cardiology 2021;77:2923–2935. doi:10.1016/j.jacc.2021.04.028.

14. Van Rooij E, Olson EN. MicroRNAs: powerful new regulators of heart disease and provocative therapeutic targets. J Clin Invest 2007;117:2369–2376. doi:10.1172/JCI33099.

15. Van Rooij E, Sutherland LB, Qi X, Richardson JA, Hill J, Olson EN. Control of Stress-Dependent Cardiac Growth and Gene Expression by a MicroRNA. Science 2007;316:575–579. doi:10.1126/science.1139089.

16. Yuan T, Krishnan J. Non-coding RNAs in Cardiac Regeneration. Front Physiol 2021;12:650566. doi:10.3389/fphys.2021.650566.

17. Lewis BP, Burge CB, Bartel DP. Conserved Seed Pairing, Often Flanked by Adenosines, Indicates that Thousands of Human Genes are MicroRNA Targets. Cell 2005;120:15–20. doi:10.1016/j.cell.2004.12.035.

18. Wilson JA, Richardson CD. Hepatitis C Virus Replicons Escape RNA Interference Induced by a Short Interfering RNA Directed against the NS5b Coding Region. J Virol 2005;79:7050–7058. doi:10.1128/JVI.79.11.7050-7058.2005.

19. Shih Y-M, Sun C-P, Chou H-H, Wu T-H, Chen C-C, Wu P-Y, et al. Combinatorial RNA Interference Therapy Prevents Selection of Pre-existing HBV Variants in Human Liver Chimeric Mice. Sci Rep 2015;5:15259. doi:10.1038/srep15259.

20. Grimm D, Kay MA. Combinatorial RNAi: A Winning Strategy for the Race Against Evolving Targets? Molecular Therapy 2007;15:878–888. doi:10.1038/sj.mt.6300116.

21. Huang Z, Shi J, Gao Y, Cui C, Zhang S, Li J, et al. HMDD v3.0: a database for experimentally supported human microRNA–disease associations. Nucleic Acids Research 2019;47:D1013–D1017. doi:10.1093/nar/gky1010.

22. Wagner JUG, Pham MD, Nicin L, Hammer M, Bottermann K, Yuan T, et al. Dissection of heterocellular cross-talk in vascularized cardiac tissue mimetics. Journal of Molecular and Cellular Cardiology 2020;138:269–282. doi:10.1016/j.yjmcc.2019.12.005.

23. Krishnan J, Suter M, Windak R, Krebs T, Felley A, Montessuit C, et al. Activation of a HIF1α-PPARγ Axis Underlies the Integration of Glycolytic and Lipid Anabolic Pathways in Pathologic Cardiac Hypertrophy. Cell Metabolism 2009;9:512–524. doi:10.1016/j.cmet.2009.05.005.

24. Kim D, Paggi JM, Park C, Bennett C, Salzberg SL. Graph-based genome alignment and genotyping with HISAT2 and HISAT-genotype. Nat Biotechnol 2019;37:907–915. doi:10.1038/s41587-019-0201-4.

25. Love MI, Huber W, Anders S. Moderated estimation of fold change and dispersion for RNA-seq data with DESeq2. Genome Biol 2014;15:550. doi:10.1186/s13059-014-0550-8.

26. Hodgkinson CP, Kang MH, Dal-Pra S, Mirotsou M, Dzau VJ. MicroRNAs and Cardiac Regeneration. Circ Res 2015;116:1700–1711. doi:10.1161/CIRCRESAHA.116.304377.

27. Richardson GD. Simultaneous Assessment of Cardiomyocyte DNA Synthesis and Ploidy: A Method to Assist Quantification of Cardiomyocyte Regeneration and Turnover. JoVE 2016:53979. doi:10.3791/53979.

28. Porrello ER, Mahmoud AI, Simpson E, Hill JA, Richardson JA, Olson EN, et al. Transient Regenerative Potential of the Neonatal Mouse Heart. Science 2011;331:1078– 1080. doi:10.1126/science.1200708.

29. Ibanez B, James S, Agewall S, Antunes MJ, Bucciarelli-Ducci C, Bueno H, et al. 2017 ESC Guidelines for the management of acute myocardial infarction in patients presenting with ST-segment elevation. European Heart Journal 2018;39:119–177. doi:10.1093/eurheartj/ehx393.

30. Thygesen K, Alpert JS, Jaffe AS, Chaitman BR, Bax JJ, Morrow DA, et al. Fourth Universal Definition of Myocardial Infarction (2018). Circulation 2018;138. doi:10.1161/CIR.0000000000000617.

31. Courtoy GE, Leclercq I, Froidure A, Schiano G, Morelle J, Devuyst O, et al. Digital Image Analysis of Picrosirius Red Staining: A Robust Method for Multi-Organ Fibrosis Quantification and Characterization. Biomolecules 2020;10:1585. doi:10.3390/biom10111585.

32. Menendez-Montes I, Garry DJ, Zhang J (Jay), Sadek HA. Metabolic Control of Cardiomyocyte Cell Cycle. Methodist DeBakey Cardiovascular Journal 2023;19:26–36. doi:10.14797/mdcvj.1309.

33. Cardoso AC, Lam NT, Savla JJ, Nakada Y, Pereira AHM, Elnwasany A, et al. Mitochondrial substrate utilization regulates cardiomyocyte cell-cycle progression. Nat Metab 2020;2:167–178. doi:10.1038/s42255-020-0169-x.

34. Zhu C, Baumgarten N, Wu M, Wang Y, Das AP, Kaur J, et al. CVD-associated SNPs with regulatory potential reveal novel non-coding disease genes. Hum Genomics 2023;17:69. doi:10.1186/s40246-023-00513-4.

35. Wagner JUG, Tombor LS, Malacarne PF, Kettenhausen L-M, Panthel J, Kujundzic H, et al. Aging impairs the neurovascular interface in the heart. Science 2023;381:897–906. doi:10.1126/science.ade4961.

36. Nicin L, Schroeter SM, Glaser SF, Schulze-Brüning R, Pham M-D, Hille SS, et al. A human cell atlas of the pressure-induced hypertrophic heart. Nat Cardiovasc Res 2022;1:174–185. doi:10.1038/s44161-022-00019-7.

37. Schwan J, Campbell SG. Prospects for In Vitro Myofilament Maturation in Stem Cell-Derived Cardiac Myocytes. Biomark Insights 2015;10:91–103. doi:10.4137/BMI.S23912.

38. Kirk EP, Sunde M, Costa MW, Rankin SA, Wolstein O, Castro ML, et al. Mutations in cardiac T-box factor gene TBX20 are associated with diverse cardiac pathologies, including defects of septation and valvulogenesis and cardiomyopathy. Am J Hum Genet 2007;81:280–291. doi:10.1086/519530.

39. Xiang F, Guo M, Yutzey KE. Overexpression of Tbx20 in Adult Cardiomyocytes Promotes Proliferation and Improves Cardiac Function After Myocardial Infarction. Circulation 2016;133:1081–1092. doi:10.1161/CIRCULATIONAHA.115.019357.

40. Arndt A-K, Schafer S, Drenckhahn J-D, Sabeh MK, Plovie ER, Caliebe A, et al. Fine mapping of the 1p36 deletion syndrome identifies mutation of PRDM16 as a cause of cardiomyopathy. Am J Hum Genet 2013;93:67–77. doi:10.1016/j.ajhg.2013.05.015.

41. Smith SJ, Towers N, Saldanha JW, Shang CA, Mahmood SR, Taylor WR, et al. The cardiac-restricted protein ADP-ribosylhydrolase-like 1 is essential for heart chamber outgrowth and acts on muscle actin filament assembly. Dev Biol 2016;416:373–388. doi:10.1016/j.ydbio.2016.05.006.

42. Stachowski-Doll MJ, Papadaki M, Martin TG, Ma W, Gong HM, Shao S, et al. GSK-3β Localizes to the Cardiac Z-Disc to Maintain Length Dependent Activation. Circ Res 2022;130:871–886. doi:10.1161/CIRCRESAHA.121.319491.

43. Zhang J, Lin Y, Zhang Y, Lan Y, Lin C, Moon AM, et al. *Frs2* α-deficiency in cardiac progenitors disrupts a subset of FGF signals required for outflow tract morphogenesis. Development 2008;135:3611–3622. doi:10.1242/dev.025361.

44. McLendon JM, Zhang X, Matasic DS, Kumar M, Koval OM, Grumbach IM, et al. Knockout of Sorbin And SH3 Domain Containing 2 (Sorbs2) in Cardiomyocytes Leads to Dilated Cardiomyopathy in Mice. J Am Heart Assoc 2022;11:e025687. doi:10.1161/JAHA.122.025687.

45. Lei Q-Y, Zhang H, Zhao B, Zha Z-Y, Bai F, Pei X-H, et al. TAZ promotes cell proliferation and epithelial-mesenchymal transition and is inhibited by the hippo pathway. Mol Cell Biol 2008;28:2426–2436. doi:10.1128/MCB.01874-07.

46. Schauer A, Adams V, Poitz DM, Barthel P, Joachim D, Friedrich J, et al. Loss of Sox9 in cardiomyocytes delays the onset of cardiac hypertrophy and fibrosis. Int J Cardiol 2019;282:68–75. doi:10.1016/j.ijcard.2019.01.078.

47. Geier C, Gehmlich K, Ehler E, Hassfeld S, Perrot A, Hayess K, et al. Beyond the sarcomere: CSRP3 mutations cause hypertrophic cardiomyopathy. Hum Mol Genet 2008;17:2753–2765. doi:10.1093/hmg/ddn160.

48. van Eldik W, den Adel B, Monshouwer-Kloots J, Salvatori D, Maas S, van der Made I, et al. Z-disc protein CHAPb induces cardiomyopathy and contractile dysfunction in the postnatal heart. PLoS One 2017;12:e0189139. doi:10.1371/journal.pone.0189139.

49. Malfatti E, Böhm J, Lacène E, Beuvin M, Romero NB, Laporte J. A Premature Stop Codon in MYO18B is Associated with Severe Nemaline Myopathy with Cardiomyopathy. J Neuromuscul Dis 2015;2:219–227. doi:10.3233/JND-150085.

50. Alazami AM, Kentab AY, Faqeih E, Mohamed JY, Alkhalidi H, Hijazi H, et al. A novel syndrome of Klippel-Feil anomaly, myopathy, and characteristic facies is linked to a null mutation in MYO18B. J Med Genet 2015;52:400–404. doi:10.1136/jmedgenet-2014-102964.

51. Sinn HW, Balsamo J, Lilien J, Lin JJ-C. Localization of the novel Xin protein to the adherens junction complex in cardiac and skeletal muscle during development. Dev Dyn 2002;225:1–13. doi:10.1002/dvdy.10131.

52. Cucoranu I, Clempus R, Dikalova A, Phelan PJ, Ariyan S, Dikalov S, et al. NAD(P)H oxidase 4 mediates transforming growth factor-beta1-induced differentiation of cardiac fibroblasts into myofibroblasts. Circ Res 2005;97:900–907. doi:10.1161/01.RES.0000187457.24338.3D.

53. Antzelevitch C, Pollevick GD, Cordeiro JM, Casis O, Sanguinetti MC, Aizawa Y, et al. Loss-of-function mutations in the cardiac calcium channel underlie a new clinical entity characterized by ST-segment elevation, short QT intervals, and sudden cardiac death. Circulation 2007;115:442–449. doi:10.1161/CIRCULATIONAHA.106.668392.

54. Kasahara A, Cipolat S, Chen Y, Dorn GW, Scorrano L. Mitochondrial fusion directs cardiomyocyte differentiation via calcineurin and Notch signaling. Science 2013;342:734–737. doi:10.1126/science.1241359.

55. Mohler PJ, Schott J-J, Gramolini AO, Dilly KW, Guatimosim S, duBell WH, et al. Ankyrin-B mutation causes type 4 long-QT cardiac arrhythmia and sudden cardiac death. Nature 2003;421:634–639. doi:10.1038/nature01335.

56. Guy PM, Kenny DA, Gill GN. The PDZ domain of the LIM protein enigma binds to beta-tropomyosin. Mol Biol Cell 1999;10:1973–1984. doi:10.1091/mbc.10.6.1973.

57. Sun B, Yao J, Ni M, Wei J, Zhong X, Guo W, et al. Cardiac ryanodine receptor calcium release deficiency syndrome. Sci Transl Med 2021;13:eaba7287. doi:10.1126/scitranslmed.aba7287.

58. Wigle JT, Harvey N, Detmar M, Lagutina I, Grosveld G, Gunn MD, et al. An essential role for Prox1 in the induction of the lymphatic endothelial cell phenotype. EMBO J 2002;21:1505–1513. doi:10.1093/emboj/21.7.1505.

59. Chen H, Chédotal A, He Z, Goodman CS, Tessier-Lavigne M. Neuropilin-2, a novel member of the neuropilin family, is a high affinity receptor for the semaphorins Sema E and Sema IV but not Sema III. Neuron 1997;19:547–559. doi:10.1016/s0896-6273(00)80371-2.

60. Azhar M, Schultz JEJ, Grupp I, Dorn GW, Meneton P, Molin DGM, et al. Transforming growth factor beta in cardiovascular development and function. Cytokine Growth Factor Rev 2003;14:391–407. doi:10.1016/s1359-6101(03)00044-3.

61. Nie Z, Xue Y, Yang D, Zhou S, Deroo BJ, Archer TK, et al. A specificity and targeting subunit of a human SWI/SNF family-related chromatin-remodeling complex. Mol Cell Biol 2000;20:8879–8888. doi:10.1128/MCB.20.23.8879-8888.2000.

62. Wang Z, Zhai W, Richardson JA, Olson EN, Meneses JJ, Firpo MT, et al. Polybromo protein BAF180 functions in mammalian cardiac chamber maturation. Genes Dev 2004;18:3106–3116. doi:10.1101/gad.1238104.

63. Hall DD, Spitler KM, Grueter CE. Disruption of cardiac Med1 inhibits RNA polymerase II promoter occupancy and promotes chromatin remodeling. Am J Physiol Heart Circ Physiol 2019;316:H314–H325. doi:10.1152/ajpheart.00580.2018.

64. Hong S, Cho Y-W, Yu L-R, Yu H, Veenstra TD, Ge K. Identification of JmjC domain-containing UTX and JMJD3 as histone H3 lysine 27 demethylases. Proc Natl Acad Sci U S A 2007;104:18439–18444. doi:10.1073/pnas.0707292104.

65. Sharma M, Li X, Wang Y, Zarnegar M, Huang C-Y, Palvimo JJ, et al. hZimp10 is an androgen receptor co-activator and forms a complex with SUMO-1 at replication foci. EMBO J 2003;22:6101–6114. doi:10.1093/emboj/cdg585.

66. Etheridge SL, Ray S, Li S, Hamblet NS, Lijam N, Tsang M, et al. Murine dishevelled 3 functions in redundant pathways with dishevelled 1 and 2 in normal cardiac outflow tract, cochlea, and neural tube development. PLoS Genet 2008;4:e1000259. doi:10.1371/journal.pgen.1000259.

67. Li X, Wu F, Günther S, Looso M, Kuenne C, Zhang T, et al. Inhibition of fatty acid oxidation enables heart regeneration in adult mice. Nature 2023;622:619–626. doi:10.1038/s41586-023-06585-5.

68. Miklas JW, Levy S, Hofsteen P, Mex DI, Clark E, Muster J, et al. Amino acid primed mTOR activity is essential for heart regeneration. iScience 2022;25:103574. doi:10.1016/j.isci.2021.103574.

69. Sakabe M, Thompson M, Chen N, Verba M, Hassan A, Lu R, et al. Inhibition of β1- AR/Gαs signaling promotes cardiomyocyte proliferation in juvenile mice through activation of RhoA-YAP axis. eLife 2022;11:e74576. doi:10.7554/eLife.74576.

70. Mohamed TMA, Abouleisa R, Hill BG. Metabolic Determinants of Cardiomyocyte Proliferation. Stem Cells 2022;40:458–467. doi:10.1093/stmcls/sxac016.

71. Huang L, Wang Q, Gu S, Cao N. Integrated metabolic and epigenetic mechanisms in cardiomyocyte proliferation. Journal of Molecular and Cellular Cardiology 2023;181:79–88. doi:10.1016/j.yjmcc.2023.06.002.

72. Clémot M, Sênos Demarco R, Jones DL. Lipid Mediated Regulation of Adult Stem Cell Behavior. Front Cell Dev Biol 2020;8:115. doi:10.3389/fcell.2020.00115.

73. Chong D, Gu Y, Zhang T, Xu Y, Bu D, Chen Z, et al. Neonatal ketone body elevation regulates postnatal heart development by promoting cardiomyocyte mitochondrial maturation and metabolic reprogramming. Cell Discov 2022;8:106. doi:10.1038/s41421-022-00447-6.

74. Talman V, Teppo J, Pöhö P, Movahedi P, Vaikkinen A, Karhu ST, et al. Molecular Atlas of Postnatal Mouse Heart Development. JAHA 2018;7:e010378. doi:10.1161/JAHA.118.010378.

75. Rasool A, Zulfajri M, Gulzar A, Hanafiah MM, Unnisa SA, Mahboob M. In vitro effects of cobalt nanoparticles on aspartate aminotransferase and alanine aminotransferase activities of wistar rats. Biotechnol Rep (Amst) 2020;26:e00453. doi:10.1016/j.btre.2020.e00453.

76. Fluiter K, ten Asbroek ALMA, de Wissel MB, Jakobs ME, Wissenbach M, Olsson H, et al. In vivo tumor growth inhibition and biodistribution studies of locked nucleic acid (LNA) antisense oligonucleotides. Nucleic Acids Res 2003;31:953–962. doi:10.1093/nar/gkg185.

77. Wang G, Rao N, Liu D, Jiang H, Liu K, Yang F, et al. Analysis of potential roles of combinatorial microRNA regulation in occurrence of valvular heart disease with atrial fibrillation based on computational evidences. PLoS ONE 2019;14:e0221900. doi:10.1371/journal.pone.0221900.

78. Xu J, Li C-X, Li Y-S, Lv J-Y, Ma Y, Shao T-T, et al. MiRNA–miRNA synergistic network: construction via co-regulating functional modules and disease miRNA topological features. Nucleic Acids Research 2011;39:825–836. doi:10.1093/nar/gkq832.

79. Mohamed TMA, Ang Y-S, Radzinsky E, Zhou P, Huang Y, Elfenbein A, et al. Regulation of Cell Cycle to Stimulate Adult Cardiomyocyte Proliferation and Cardiac Regeneration. Cell 2018;173:104–116.e12. doi:10.1016/j.cell.2018.02.014.

80. Chen Y, Lüttmann FF, Schoger E, Schöler HR, Zelarayán LC, Kim K-P, et al. Reversible reprogramming of cardiomyocytes to a fetal state drives heart regeneration in mice. Science 2021;373:1537–1540. doi:10.1126/science.abg5159.

81. Hashmi S, Ahmad HR. Molecular switch model for cardiomyocyte proliferation. Cell Regeneration 2019;8:12–20. doi:10.1016/j.cr.2018.11.002.

82. Tian Y, Liu Y, Wang T, Zhou N, Kong J, Chen L, et al. A microRNA-Hippo pathway that promotes cardiomyocyte proliferation and cardiac regeneration in mice. Sci Transl Med 2015;7. doi:10.1126/scitranslmed.3010841.

83. Ziff OJ, Bromage DI, Yellon DM, Davidson SM. Therapeutic strategies utilizing SDF-1α in ischaemic cardiomyopathy. Cardiovascular Research 2018;114:358–367. doi:10.1093/cvr/cvx203.

84. Hullinger TG, Montgomery RL, Seto AG, Dickinson BA, Semus HM, Lynch JM, et al. Inhibition of miR-15 Protects Against Cardiac Ischemic Injury. Circ Res 2012;110:71–81. doi:10.1161/CIRCRESAHA.111.244442.

85. Zhao Y, Ransom JF, Li A, Vedantham V, Von Drehle M, Muth AN, et al. Dysregulation of Cardiogenesis, Cardiac Conduction, and Cell Cycle in Mice Lacking miRNA-1-2. Cell 2007;129:303–317. doi:10.1016/j.cell.2007.03.030.

86. Zhao Y, Samal E, Srivastava D. Serum response factor regulates a muscle-specific microRNA that targets Hand2 during cardiogenesis. Nature 2005;436:214–220. doi:10.1038/nature03817.

87. Gan J, Tang FMK, Su X, Lu G, Xu J, Lee HSS, et al. microRNA-1 inhibits cardiomyocyte proliferation in mouse neonatal hearts by repressing CCND1 expression. Ann Transl Med 2019;7:455–455. doi:10.21037/atm.2019.08.68.

88. Hao Y-L, Fang H-C, Zhao H-L, Li X-L, Luo Y, Wu B-Q, et al. The role of microRNA-1 targeting of *MAPK3* in myocardial ischemia-reperfusion injury in rats undergoing sevoflurane preconditioning via the PI3K/Akt pathway. American Journal of Physiology-Cell Physiology 2018;315:C380–C388. doi:10.1152/ajpcell.00310.2017.

89. Pan Z, Sun X, Ren J, Li X, Gao X, Lu C, et al. miR-1 Exacerbates Cardiac Ischemia-Reperfusion Injury in Mouse Models. PLoS ONE 2012;7:e50515. doi:10.1371/journal.pone.0050515.

90. Lagos-Quintana M, Rauhut R, Yalcin A, Meyer J, Lendeckel W, Tuschl T. Identification of Tissue-Specific MicroRNAs from Mouse. Current Biology 2002;12:735–739. doi:10.1016/S0960-9822(02)00809-6.

91. Lee RC, Ambros V. An Extensive Class of Small RNAs in *Caenorhabditis elegans*. Science 2001;294:862–864. doi:10.1126/science.1065329.

92. Foinquinos A, Batkai S, Genschel C, Viereck J, Rump S, Gyöngyösi M, et al. Preclinical development of a miR-132 inhibitor for heart failure treatment. Nat Commun 2020;11:633. doi:10.1038/s41467-020-14349-2.

93. Abplanalp WT, Fischer A, John D, Zeiher AM, Gosgnach W, Darville H, et al. Efficiency and Target Derepression of Anti-miR-92a: Results of a First in Human Study. Nucleic Acid Therapeutics 2020;30:335–345. doi:10.1089/nat.2020.0871.

94. Kim T, Croce CM. MicroRNA: trends in clinical trials of cancer diagnosis and therapy strategies. Exp Mol Med 2023;55:1314–1321. doi:10.1038/s12276-023-01050-9.

95. Hinkel R, Ramanujam D, Kaczmarek V, Howe A, Klett K, Beck C, et al. AntimiR-21 Prevents Myocardial Dysfunction in a Pig Model of Ischemia/Reperfusion Injury. Journal of the American College of Cardiology 2020;75:1788–1800. doi:10.1016/j.jacc.2020.02.041.

